# Mechanistic modeling of brain metastases in NSCLC provides computational markers for personalized prediction of outcome

**DOI:** 10.1101/2023.01.10.23284189

**Authors:** Sébastien Benzekry, Pirmin Schlicke, Pascale Tomasini, Eléonore Simon

**Author notes:** Corresponding author, Faculté de Pharmacie, 27 boulevard Jean Moulin, 13005 Marseille, France. These authors contributed equally and are co-first authors in alphabetical order (Sébastien Benzekry, Pirmin Schlicke). Social media handles. Authors: @SBenzekry, @PirminSchlicke. Institutions: @aphm_actu, @crcm_marseille, @inria_sophia, @tu_muenchen.

## Abstract

**Background:** Intracranial progression after curative treatment of early-stage non-small cell lung cancer (NSCLC) occurs from 10 to 50% and is difficult to manage, given the heterogeneity of clinical presentations and the variability of treatments available.

The objective of this study was to develop a mechanistic model of intracranial progression to predict survival following a first brain metastasis (BM) event.

**Methods:** Data included early-stage NSCLC patients treated with a curative intent who had a BM as the first and single relapse site (N=31).

We propose a mechanistic mathematical model to estimate the amount and sizes of (visible and invisible) BMs. The two key parameters of the model are *α*, the proliferation rate of a single tumor cell; and *μ*, the per day, per cell, probability to metastasize. The predictive value of these individual computational biomarkers was evaluated.

**Findings:** The model was able to correctly describe the number and size of metastases at the time of first BM relapse for 20 patients. Parameters *α* and *μ* were significantly associated with overall survival (OS) (HR 1.65 (1.07-2.53) p=0.0029 and HR 1.95 (1.31-2.91) p=0.0109, respectively). Adding the computational markers to the clinical ones significantly improved the predictive value of OS (c-index increased from 0.585 (95% CI 0.569-0.602) to 0.713 (95% CI 0.700-0.726), p<0.0001).

**Interpretation:** We demonstrated that our model was applicable to brain oligoprogressive patients in NSCLC and that the resulting computational markers had predictive potential. This may help lung cancer physicians to guide and personalize the management of NSCLC patients with intracranial oligoprogression.

**SIGNIFICANCE STATEMENT:** Non-small cell lung cancer is difficult to manage when brain metastases are present. This study presents a mathematical model that can be calibrated on individual patients’ data early in the treatment course to explain the growth dynamics of brain metastases and demonstrates that the mathematically derived parameters can serve as predictive tool in clinical routine care.

**Highlights:**

- Mechanistic mathematical modeling allows individualized prognosis for lung cancer patients at first brain metastatic relapse
- Individual model-derived computational parameters identifies high-risk patients in terms of brain metastasis progression and survival
- Prognostic features include quantification of the number and sizes of both clinically visible and invisible brain metastases

## INTRODUCTION

The 5 years survival rate in non-small cell lung cancer (NSCLC) is only 17%^1,2^. This poor prognosis is partly due to the frequent occurrence of brain metastases (BM), occurring in up to 50% of NSCLC patients^3^ and directly responsible for death in 50% cases^4,5^. Even in initially early-stage NSCLC treated with curative intent, intracranial relapse occurs in 10% of the cases in stage I-II NSCLC and 30 to 50% cases in stage III NSCLC^6^. Not only is the brain one of the first sites of recurrence after surgery or curative radiotherapy, but it can also be the only site of relapse, then called intracranial oligoprogression^7–9^.

Oligoprogression is defined as the progression of a limited number of metastases, ranging from 3 to 5, and possibly treated by local therapies^10–12^. Existing data on intracranial oligoprogression are mostly related to palliative situations, and to patients receiving tyrosine kinase inhibitors^13^. There are no real guidelines for the management of single intracranial oligoprogression in patients with early-stage NSCLC at diagnosis, initially treated with curative intent. Furthermore, the difficulty in establishing guidelines relies on the heterogeneity of this situation. Some patients undergo a single intracranial recurrence, while others suffer from either multiple intracranial relapse or subsequent extra-cranial relapse^14^.

Several options are available in the treatment of BM, including local and systemic therapies. Local treatments include neurosurgery, stereotaxic radiosurgery (SRS) and whole brain radiotherapy (WBRT)^15–19^. Local treatment indications have evolved over time and the place of SRS is becoming increasingly important^16–20^. Nevertheless, WBRT has the advantage of covering all macroscopic and microscopic BM which would not be detected with MRI and could have a prophylactic role on the development of BM^21,22^. However, WBRT is also responsible for neurocognitive decline, leading to a lower quality of life^23,24^. On the other hand, systemic treatments include chemotherapy, targeted therapies or immunotherapy^10,25–28^. The intracranial efficacy of systemic treatments is heterogeneous and partly depends on the blood-brain barrier (BBB) crossing of the drugs^29^. Moreover, the unique brain microenvironment likely plays a critical role in treatment response, especially with immunotherapy^30–33^. The combination of systemic treatments such as chemotherapy and immunotherapy seems to induce better intracranial response than immunotherapy alone^34,35^. Combinations of local and systemic treatments also seem to have a synergistic effect and lead to better intracranial response^36^. Radiotherapy (either SRS or WBRT) could help damage the BBB, allowing the crossing of systemic treatments.

The complexity of multimodal management of BM relies on the diversity of possible treatments (including local and systemic treatments), the difficulty of crossing the BBB for systemic treatments, the specificity of the brain microenvironment and the lack of predictability of the appearance of BM, as well as further extracranial progression, as evidenced by Levy’s study of European practices^37^.

To guide the management of patients with BM, prognostic scores have been established, such as the mol-Lung GPA^38–40^. However, these scores do not help predict the occurrence of metastases, even if some predictive factors were identified such as age ≤ 62 years, stage T4 or N2-3, adenocarcinoma subtype, presence of leukoaraiosis, and molecular profile^41,42^.

Predicting the course and the appearance of BM would improve the monitoring of patients and offer a more personalized treatment. For this purpose, the contribution of mathematical modeling can be crucial.

The first mathematical models of metastatic occurrence based on experimental observations were established by Liotta in the 1970’s^43^. Further mathematical models were then established in animal models^44–46^. These models were based on two main phenomena: growth and spread of a tumor. The latter is often successfully described using the Gompertz model^47–49^. For metastatic spread, we have developed models based on pioneering work by Iwata et al^45,46,50,51^. Our team further developed these mathematical constructs to be applicable in the clinic, for example on breast^52^ or kidney^53^ cancer. This model was then developed within the framework of NSCLC BMs^54^. This last work proposed a method to determine the age of the disease according to the pathology and size of the primary tumor, to test different dissemination and colonization scenarios and finally to predict the appearance and growth of BMs over time. However, this preliminary work only included two patients.

The objective of this study was to develop the mathematical model and apply it to a situation of intracranial oligoprogression in multiple patients with early-stage NSCLC, initially treated with a curative intent. The secondary objective was to assess the association of this model with overall survival and progression-free survival.

## MATERIALS & METHODS

### Data

We retrospectively investigated all early-stage NSCLC patients treated with a curative intent who had a brain relapse at Public Assistance from Marseille Hospitals (APHM) in the department of Multidisciplinary Oncology and Therapeutic Innovations. We enrolled all patients with BM as the first relapse site. Primary tumor progression or metastatic extracranial progression at the time of the brain oligo-progression were excluded. Patients with ongoing systemic treatment at the time of oligo-progression were also excluded. This study was approved by the institution Data Protection Officer (# 2021-25).

Patients’ characteristics (including sex, age, smoking history, performance status), NSCLC data (histology, stage at diagnosis, molecular profile), treatment data (treatment received for the primary tumor, history of BM treatment) and outcome data (size and number of BM at first relapse, longitudinal data of BM, progression-free survival (PFS) and overall survival (OS)) were investigated. OS is the time from diagnosis of the primary tumor to death from any cause (or date of last follow up). PFS was defined as the time between the time point of first metastatic relapse and second metastatic relapse.

Since the model presented in this study is based on numbers of cells, the longitudinal diameter measurements of the data set were converted by assuming spherical shape calculating the volume as 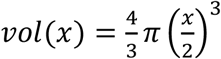 where *x* is the individual diameter value. These volumetric data were then rescaled into cell numbers using the conversion rule 1mm^3^ = 10^6^ cells^55^.

### Model definition

A primary tumor with size *S*_*P*_(*t*) at time *t* ≥ 0 was assumed to follow Gompertzian growth^56,57^. Since patients underwent surgery, we adjusted the growth equation to account for full tumor removal at day of surgery *T*_*s*_ > 0 such that

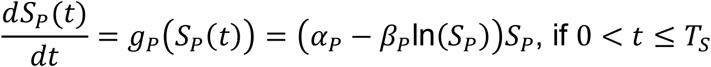

but *S*_*P*_(*t*) = 0 if *t* > *T*_*S*_. The initial size of a tumor at time *t* = 0 was assumed to be of exactly one cell, i.e., *S*_*P*_(0) = 1^58^. The parameter *α*_*P*_ can be interpreted as the initial specific growth rate of a single tumor cell and the parameter *β*_*P*_ is the exponential decay factor of this specific growth rate^59^. The analytical solution of the growth dynamics for the primary tumor and the analogous dynamics for the metastases are given in the supplementary material.

Two additional parameters *μ* and *γ* were introduced for the dissemination process: parameter *μ* stands for the probability per day and per cell for a tumor cell to succeed going through all steps of the postulated metastatic cascade^60^. Parameter *γ* ∈ [0,1] describes how the geometrical structure of the tumor enables the tumor to seed new metastases. The dissemination rate (number of newly created metastases per day) was therefore assumed to be:

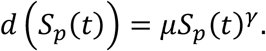

The model equations for the number of metastases (visible and invisible) are described in the supplementary material and details to this approach are given in^60^. Since there is evidence that secondary and thus lately seeded metastases are of minor clinical relevance^61^, for the purpose of our predictive framework we focused on metastases seeded by the primary tumor only.

### Parameter assumptions

Following Bilous et al.^60^ we reduced further the number of free parameters to ensure practical identifiability. We assumed a maximum carrying capacity of *exp*(*α*/*β*) = 10^12^ cells^62^ for both the primary tumor and metastases’ growth kinetics. Since longitudinal pre-diagnosis measurements of primary tumors were not available, we assumed a pathology-dependent doubling time estimated from a meta-analysis of the literature^60^. Specifically, doubling times were assumed to be 201, 104 and 91 days for adenocarcinoma, squamous cell carcinoma and undifferentiated carcinoma, respectively^60^. The metastases observed clinically, i.e., the visible BMs, are assumed to be larger than the volume corresponding to a diameter measurement of three mm. The corresponding volumetric measure is denoted by the threshold size *s*_*v*_.

### Parameter identification

The procedure to determine the size and number of BMs from the model equations and values of the parameters is described in the supplementary material. We solely used data up to the time of first metastatic relapse – denoted *T*_*BM*_ – and consequently identified the parameters using only one time point with BMs measurements. Preliminary explorations of fitting procedures showed that the parameter identification was very instable due to the unknown metastatic density distribution below the threshold size (*s*_*v*_). We therefore considered two possible scenarios for the density distribution below *s*_*v*_: i) no additional BM, i.e., all existing BM above *s*_*v*_ have been detected successfully (‘best case’) or ii) another BM of size *s*_*v*_, i.e., one BM has been overseen (‘worst case’). Our final method was a compromise between i) and ii): we assumed one artificial BM with size *s*_*v*_ with an associated number of one-half BM. From this artificial measurement, we were able to control the important variance of the metastatic density below *s*_*v*_.

The doubling time *τ*_*P*_(*s*) of a primary tumor of size *S*_*P*_(*T*_*d*_) at time of diagnosis 0 ≤ *T*_*d*_ ≤ *T*_*S*_ was used to determine the two growth parameters *α*_*P*_ and *β*_*P*_ (see supplementary material). Preliminary exploration of the parameter space suggested that a value of *γ* = 0.1 was able to describe the data. To ensure practical identifiability of the model parameters we therefore fixed *γ* = 0.1 and only considered parameters *α* and *μ* as variables during model fitting. The model was implemented in Matlab. A least squares calculation comparing model evaluations against the empirical data of the cumulative metastatic density distribution at the time of first metastatic relapse was performed using the GlobalSearch environment^63,64^, fitting for the two parameters *α* and *μ* on biologically reasonable intervals (i.e. positive values), respectively.

### Statistical analysis

These two computational parameters *α* and *μ* were used as computational biomarkers with potential additional prognosis value than the mere clinical data. We analyzed association of these parameters with overall survival and progression-free survival using a previously introduced method^65^. Log-rank statistical tests were performed on dichotomized groups (with threshold taken at the median value of the computational biomarker)^66^. In addition, multivariable proportional hazard Cox regression models were considered, either using only the clinical covariates or the clinical covariates augmented with *α* and *μ*. These were implemented using the *lifelines* python package^67^. The predictive ability of these Cox models was determined calculating the mean value of Harrell’s c-index^68^ of one hundred replications of a three-fold cross-validation procedure. In a closer examination, only covariates below a significance level p < 0.2 were selected for multivariable analysis. To determine the statistical significance of mean c-indices between Cox models, two-sample t-tests were conducted.

## RESULTS

### Data presentation and description

We identified 31 patients who had intracranial oligoprogression after initial treatment with a curative intent for early-stage NSCLC. Of these, 10 were women and 21 were men. There were 9 squamous cell carcinomas and 22 non-squamous carcinomas, including 20 adenocarcinomas. At the time of diagnosis, 20 patients were stage I/II and 11 were stage III according to the 8th TNM classification. Number of BMs lied between one and three and the corresponding diameter sizes between 4 mm and 75 mm. Treatment received for the primary tumor, the number, and the treatment of brain metastasis at the first relapse are reported in Table 1. 17 patients had more than one intracranial recurrence. Median OS was 80.98 months (95% CI 37.56 – 98.27) and median PFS was 11.73 months (95% CI 4.93 – 27.20).

**Table 1.** Data description: Patient overviewn. PT = Primary tumor, BM = Brain metastases, SRS = stereotaxic radiosurgery, WBRT = whole brain radiotherapy

| **Data population** | **Population N 31 (%)** |
| --- | --- |
| **Sex** |  |
| Female – Male | 10 (32.3%) – 21 (67.7%) |
| **Median Age at Diagnostic** | 61 (39-82) |
| **Smoking Status** |  |
| Smoker | 11 (35.5%) |
| Former smoker | 13 (41.9%) |
| No smoker | 1 (3.2%) |
| **Stage at diagnostic** |  |
| I/II | 20 (65%) |
| III | 11 (35%) |
| **Primary tumor pathology subtype** |  |
| Non squamous cell carcinoma | 22 (71%) |
| Squamous cell carcinoma | 9 (29%) |
| **Molecular Status of PT** |  |
| EGFR | 3 |
| KRAS | 4 |
| PTEN | 1 |
| TP53 | 2 |
| STK11 | 1 |
| CTNNB1 | 1 |
| No mutation | 8 |
| Unknown | 14 |
| **Size of PT at diagnosis (diameter value)** |  |
| ≤ 25 mm | 14 (45%) |
| > 25, ≤ 50 mm | 6 (19%) |
| > 50, ≤ 75 mm | 7 (23%) |
| > 75 mm | 1 (3%) |
| Unknown | 3 (10%) |
| **Surgery of the primary tumor** |  |
| Yes | 22 (71%) |
| No | 9 (29%) |
| **Systemic treatment for the primary tumor** |  |
| Yes | 19 (61.3%) |
| No | 12 (38.7%) |
| **Median time between the diagnosis and first relapse (months)** | 13.63 (95% CI 9.03 – 18.00) |
| **Number of BM at first relapse** |  |
| 1 | 18 (58.1%) |
| 2 | 5 (16.1%) |
| 3 | 8 (25.8%) |
| **Sum of BM diameters at relapse** |  |
| ≤ 20 mm | 18 (58%) |
| > 20, ≤ 40 mm | 9 (29%) |
| > 40, ≤ 60 mm | 3 (10%) |
| > 60 mm | 1 (3%) |
| **Individual BM diameters at relapse** |  |
| ≤ 10 mm | 25 (49%) |
| > 10, ≤ 20 mm | 12 (24%) |
| > 20, ≤ 30 mm | 6 (12%) |
| > 30 mm | 8 (16%) |
| **Type of local treatment at first cerebral relapse** |  |
|  | 3 (9.7%) |
| **Surgery alone** | 4 (12.9%) |
| **Surgery + WBRT** | 7 (22.6%) |
| **Surgery + SRS** | 16 (51.6%) |
| **SRS alone** | 1 (3.2%) |
| **WBRT alone** |  |
| **Systemic treatment at first cerebral relapse** |  |
| Yes | 11 (35.5%) |
| No | 20 (64.5%) |
| **Number of cerebral relapse** |  |
| 1 | 14 (45.2%) |
| 2-3 | 13 (42%) |
| >3 | 4 (12.8%) |
| **Median time between the first and second relapse (months)** | 11.73 (95% CI 4.93 – 27.20) |

The data included values of longitudinal diameter measurements determined via computed tomography (CT) and magnetic resonance imaging (MRI) images of primary tumors and BMs as well as the time of relapse and time of death. Representative patient plots of these time series data are reported in Figure 1.

**Figure 1.**
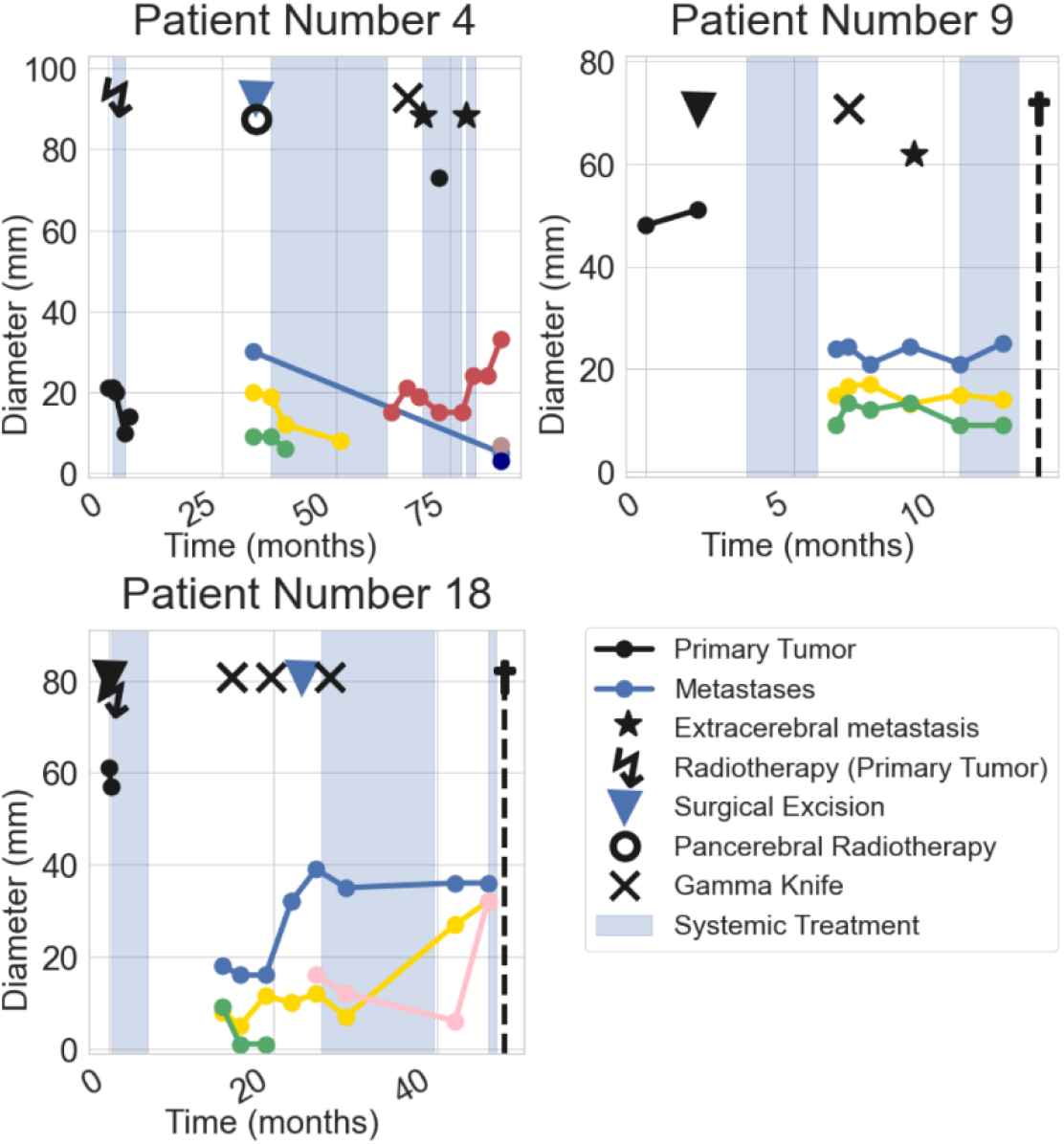
Data: Example patients. Time course of three example patients. Black bullets show the primary tumor size and colored bullets the brain metastases sizes. The time course was shifted such that time zero corresponds to the day of primary diagnosis. Also shown are local and systemic treatments (triangle, cross, flash, circle, colored background), the presence of extra-cerebral metastases (black asterisk, not linked to diameter axis) and the time of the respective patient’s death (black cross with dashed line).

Out of the 31 patients, three patients had no primary tumor measurements at primary diagnosis. In the later analysis, these patients had to be excluded, since the primary tumor growth parameter calibration could not be performed (cf. Mathematical model).

### Parameter values

As explained above, the remaining parameters subject to identification through data fitting were *α* (growth) and *μ* (dissemination). Parameter values are shown in Table 2.

**Table 2.** Parameter description. Some measures for the fitted parameter values on the population level, std = standard deviation.

| parameter | mean | median | std | min | max |
| --- | --- | --- | --- | --- | --- |
| $\alpha$ | $4.93 \cdot 10^{-2}$ | $4.03 \cdot 10^{-2}$ | $3.18 \cdot 10^{-2}$ | $1.22 \cdot 10^{-2}$ | $1.34 \cdot 10^{-1}$ |
| $\mu$ | $3.57 \cdot 10^{-4}$ | $2.33 \cdot 10^{-4}$ | $2.77 \cdot 10^{-4}$ | $8.22 \cdot 10^{-5}$ | $1.22 \cdot 10^{-3}$ |

### Simulations and outcome

The model was used to simulate history of all BMs, including those not yet clinically detectable at *T*_*BM*_. To ensure parameter identifiability, a specific method was introduced (cf. Methods). The total number of predicted BMs at time *T*_*BM*_ (clinically visible + invisible) was compared to the number of BMs detected during the full clinical history of the individual patient (Figure 2).

**Figure 2.**
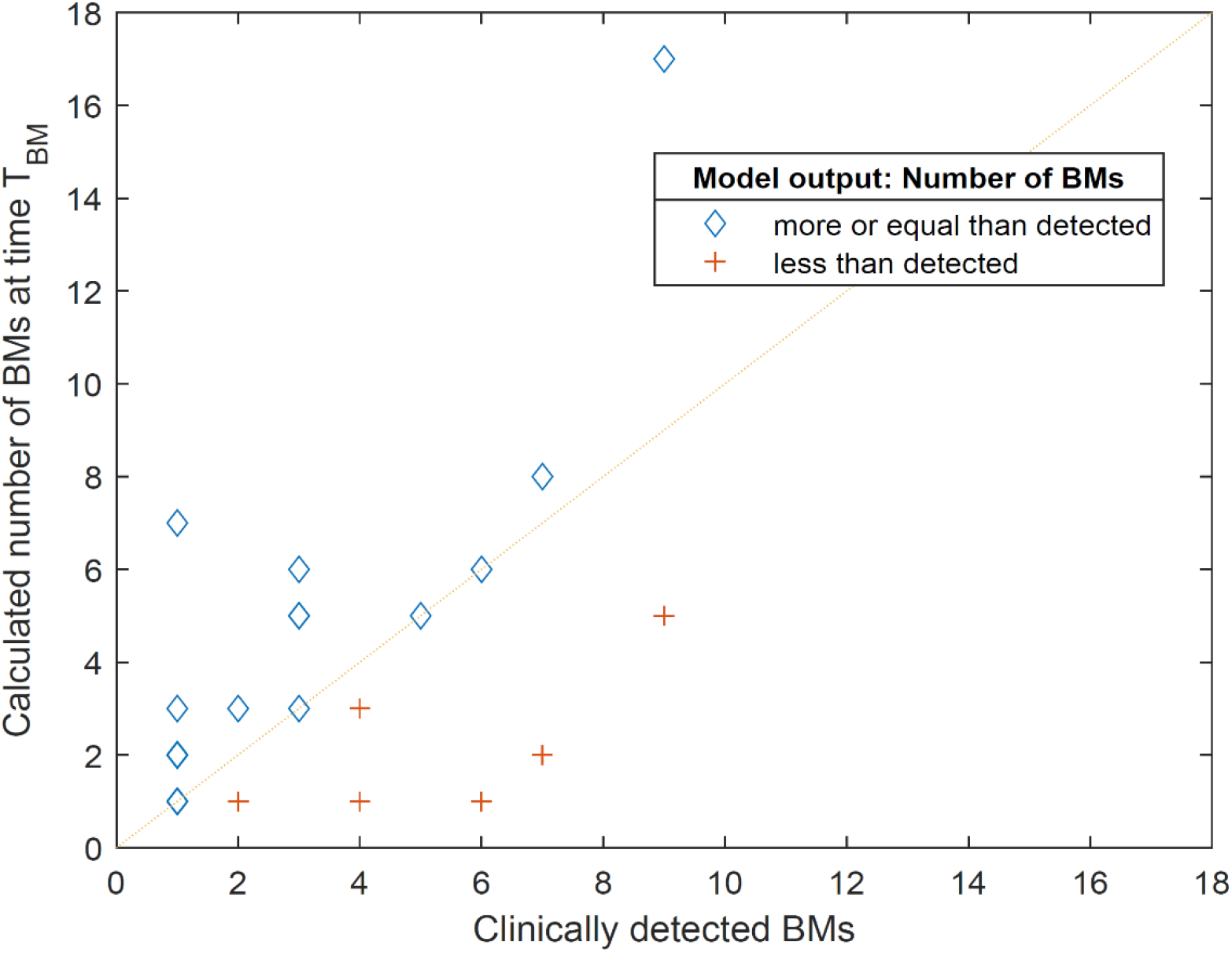
Population results for the calculated number of BMs (visible + invisible) compared to the clinically observed number over the treatment course. Estimations that prognosed less BMs than clinically observed are indicated with red crosses. Estimations that prognosed the correct or a larger number of BMs than observed are indicated as blue diamonds. Several points correspond to multiple cases: (1,1) four times, (1,2) five times, (2,1) two times, (4,1) two times and (3,5) two times.

Individual simulation of BM history for a specific patient is presented in Figure 3. Analogous simulations for all patients are shown in the supplementary material. Figure 3A shows the data of the clinical history. Panels 3B and 3C compare the model-predicted metastatic distributions at the two time points of primary diagnosis and *T*_*BM*_, respectively. The whole-time course of primary tumor and metastases sizes is presented in Figure 3D. The simulated visible metastases in comparison to the clinically detected metastases at time *T*_*BM*_ as well as the size distributions of BMs at *T*_*BM*_ are presented in Figure 3E. Compared to the data, the model calculations produced reasonable estimations of size and number of visible BMs at *T*_*BM*_ throughout the data set. For 20 patients the calculated theoretical number of BMs at time *T*_*BM*_ was greater or equal to the number of BMs observed during the full clinical treatment of the respective patient. We considered this not only as a reasonable estimate but also a good indication that the model-predicted total number and size of BMs proves to be useful in estimating the invisible BMs early in the treatment course.

**Figure 3.**
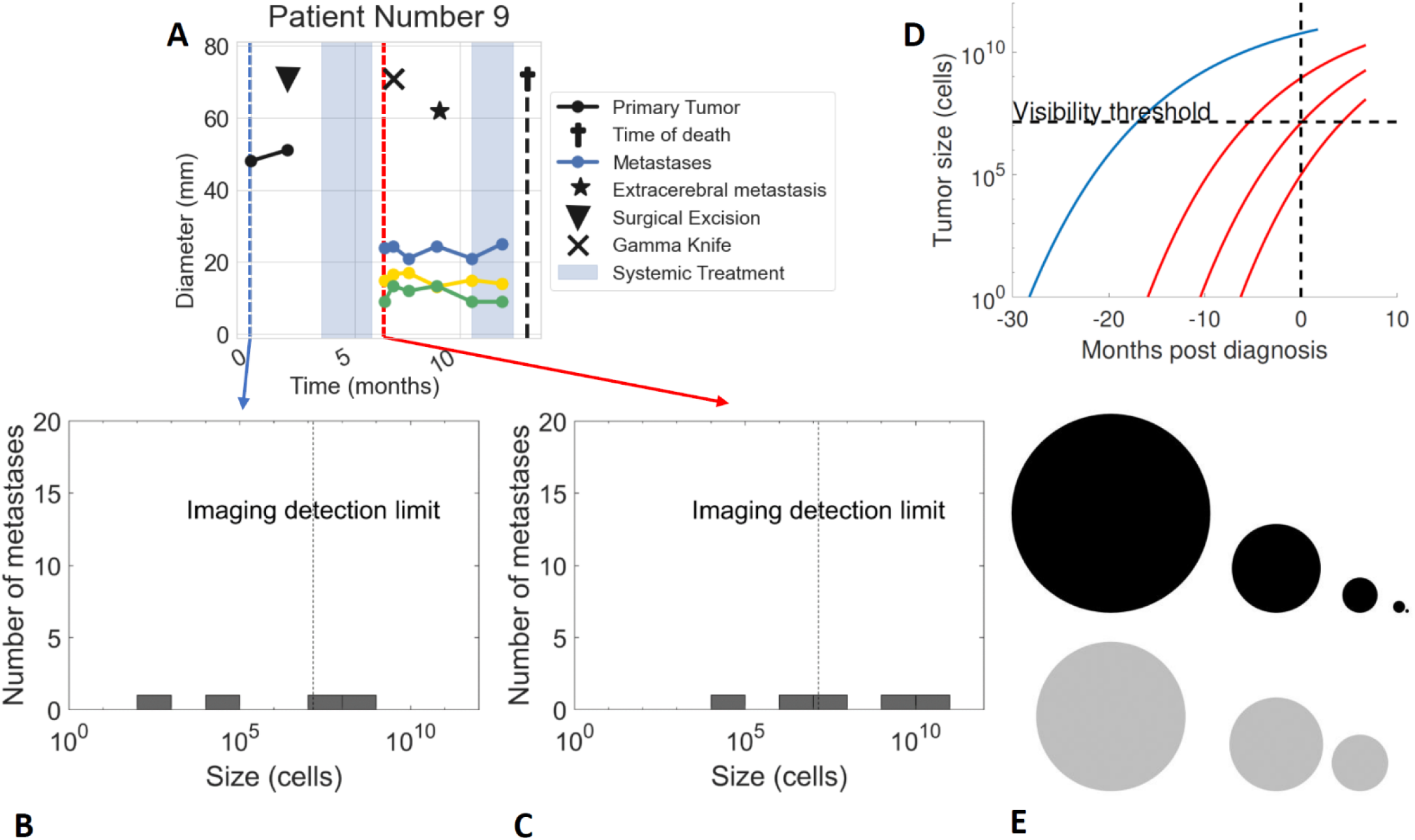
Simulation of example patient. **A**. Time series data of an example patient. Data time points used for estimation of the deterministic model parameters are the primary diagnosis (blue dashed line) and the time of first metastatic relapse T_BM_ (red dashed line). The size of the primary tumor at primary diagnosis determines the growth parameters for the primary tumor growth dynamics (cf. text). From the number and size of metastases at first metastatic relapse the two free model parameters were fitted. **B**. Model calculation allows to compute the metastatic distribution over time for the primary diagnosis. **C**. Further calculation shows the metastatic distribution at time T_BM_. Visible metastases are the ones to the right of the imaging detection limit (gray dotted line). **D**. Growth dynamics can be followed for the primary tumor (blue solid line) and the metastases (red solid line) respectively. **E**. Comparison of the computed BMs sizes at time T_BM_ (black) with the observed data (gray).

We then compared two patients with the same number of metastases at time *T*_*BM*_ and relatively familiar time frames. Interestingly, the model yielded very different profiles for the invisible BMs and therefore might suggest different treatment approaches for the two patients (Figure 4). Even though the clinical data at *T*_*BM*_ was very similar for the two patients, the model predicted different outcomes. For instance, the number of visible and invisible BMs at *T*_*BM*_ were 5 and 8, respectively. These numbers again appeared reasonable when compared to the full clinical treatment course of each patient.

**Figure 4.**
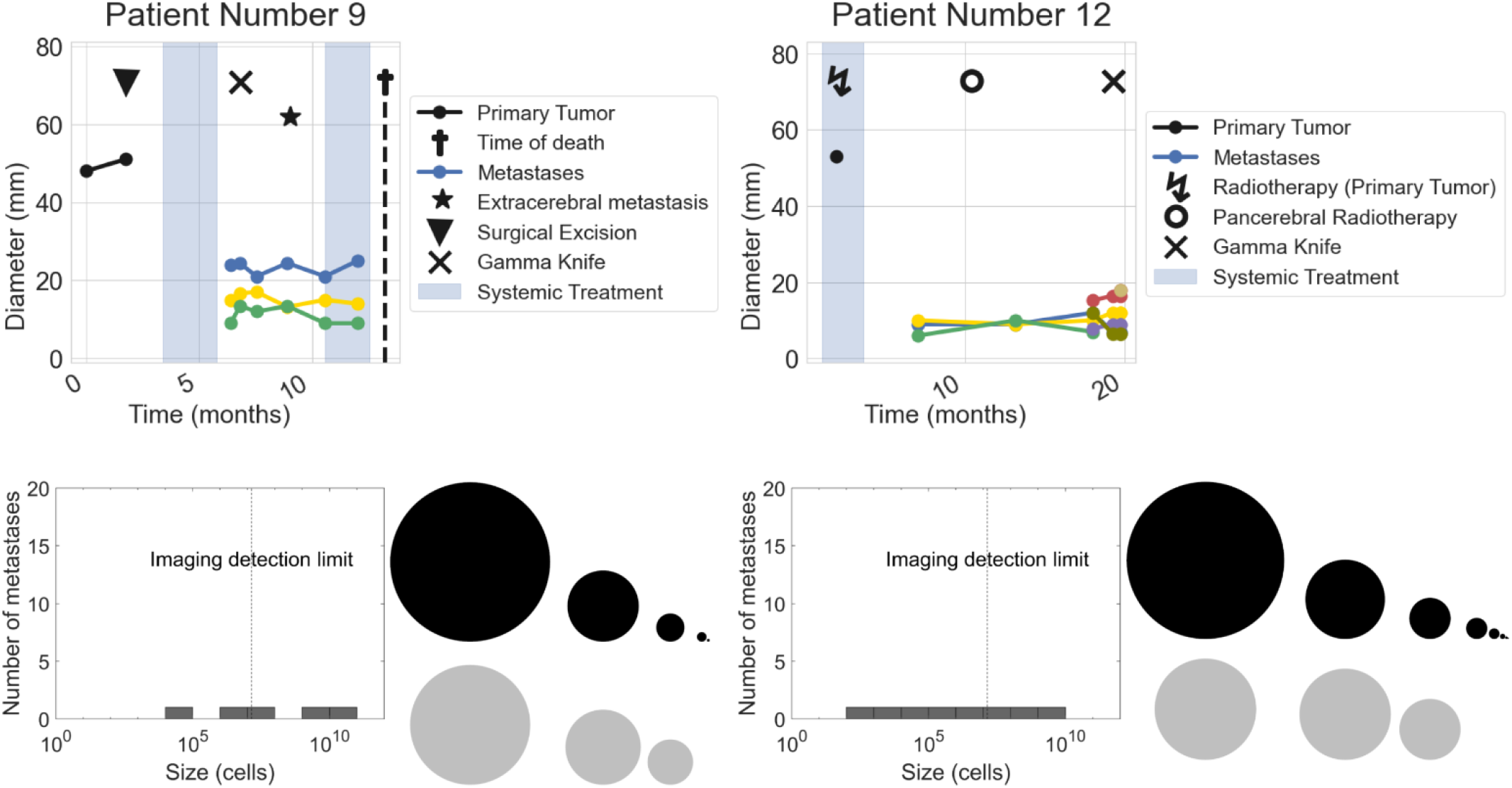
Comparison of two patients. The upper row shows the full clinical history of the two patients. The estimation of the metastatic distribution at time of first metastatic relapse and the spherical representations of the corresponding metastases’ sizes (black) are shown in the lower row for the respective patient, compared to the data of clinically detectable metastases (gray).

### Statistical evaluation and prediction models

To assess the predictive power of the computational markers *α* and *μ*, dichotomized Kaplan-Meier were generated for OS and PFS (Figure 5). For *α*, significant differences were observed for OS (*p* = 0.0026) and PFS (*p* = 0.0108). For *μ*, a significant difference was found at the 80% quantile for OS (*p* = 0.0356) and PFS (*p* = 0.0254), thus identifying a fraction of high-risk patients. We further assessed the association with outcome of *α* and *μ*, as well as the clinical variables in univariate Cox models (Table 3). Here, we found statistically significant hazard ratios for both the computationally determined markers *α* and *μ* for OS (*p* = 0.0229 and *p* = 0.0011, respectively).

**Figure 5.**
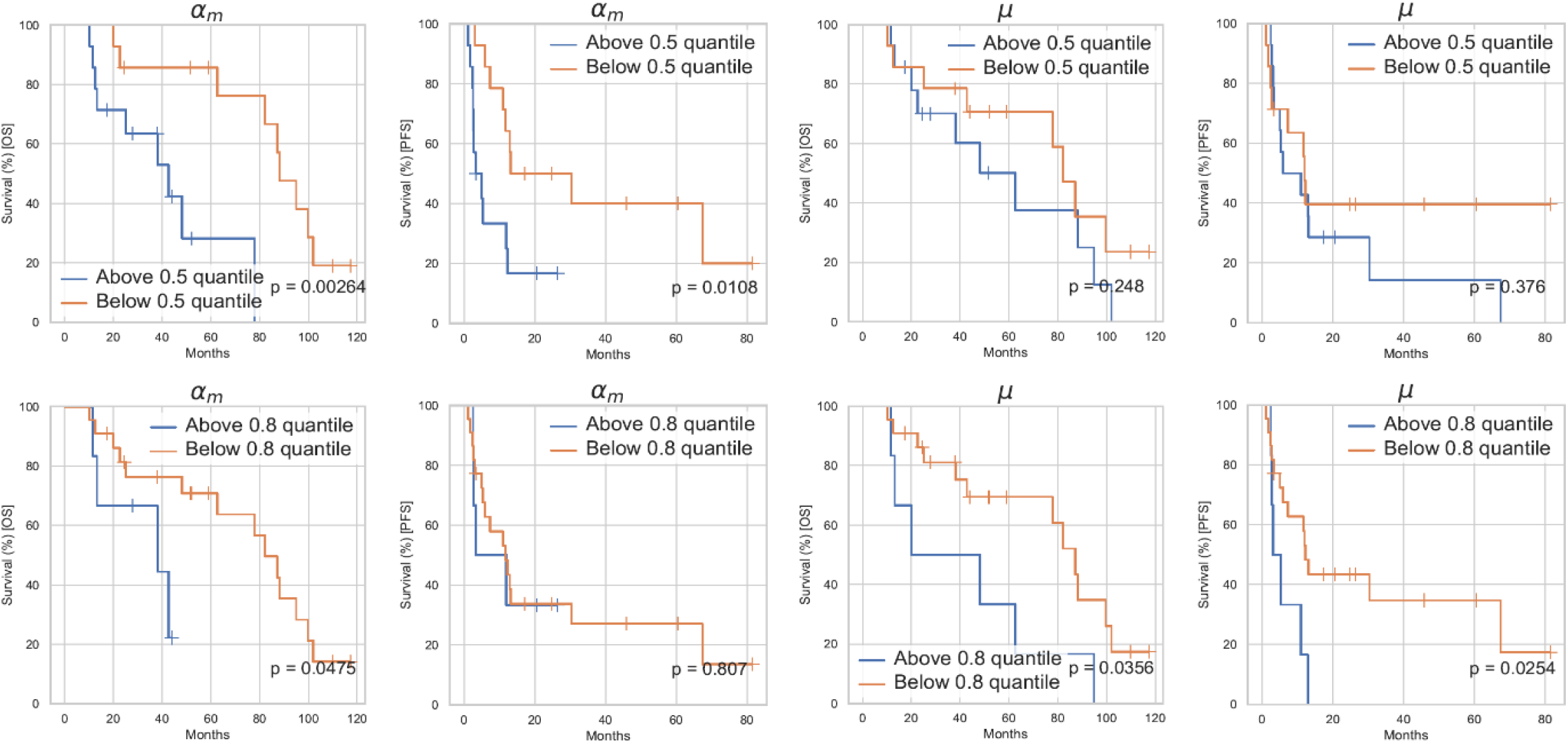
Kaplan-Meier curves for the computational parameters α and μ. The survival curves for the two computational parameter groups above and below the population’s median value (first row) and the 80% quantile value (second row) with respect to overall survival (first and third column) and progression-free survival (second and fourth column). The corresponding calculated p-value from the log-rank test for dichotomized groups is shown in the right lower corner.

**Table 3:** Results of the univariate Cox regression for OS and PFS. The table shows the characteristics of the results of the univariate Cox regression including clinical and computational covariates for overall survival and progression-free survival. HR = hazard ratio, CI = confidence interval, BM = brain metastases, PT = primary tumor.

| Covariate | Hazard Ratio (OS) | p-value (OS) | 95% CI (OS) | Hazard Ratio (PFS) | p-value (PFS) | 95% CI (PFS) |
| --- | --- | --- | --- | --- | --- | --- |
| Sex | 1.1 | 0.602 | 0.768 - 1.58 | 0.984 | 0.936 | 0.663 - 1.46 |
| Age at diagnosis | 1.03 | 0.900 | 0.679 - 1.55 | 0.738 | 0.167 | 0.479 - 1.14 |
| Number of bm at relapse | 1.72 | 0.00556 | 1.17 - 2.53 | 1.25 | 0.255 | 0.851 - 1.84 |
| Stage at diagnosis | 0.866 | 0.435 | 0.604 - 1.24 | 1.1 | 0.674 | 0.716 - 1.67 |
| Histology of pt | 0.792 | 0.223 | 0.545 - 1.15 | 0.715 | 0.117 | 0.47 - 1.09 |
| Parameter $\alpha$ | 1.65 | 0.0229 | 1.07 - 2.53 | 1.25 | 0.237 | 0.862 - 1.83 |
| Parameter $\mu$ | 1.95 | 0.00109 | 1.31 - 2.91 | 1.54 | 0.0729 | 0.961 - 2.46 |
| Size of PT at diagnosis | 1.05 | 0.781 | 0.755 - 1.45 | 0.9 | 0.648 | 0.574 - 1.41 |
| Size of BM at relapse | 0.992 | 0.967 | 0.677 - 1.45 | 1.52 | 0.0396 | 1.02 - 2.26 |
| Whole-brain radiotherapy after $T_{BM}$ | 0.89 | 0.539 | 0.615 - 1.29 | 0.649 | 0.0282 | 0.442 - 0.955 |
| Systemic treatment after $T_{BM}$ | 1.16 | 0.420 | 0.807 - 1.67 | 0.824 | 0.359 | 0.546 - 1.25 |

To determine the added value of the computational markers in comparison to the clinical data, we tested their significance in multivariable Cox models built on the clinical data alone or the clinical data and the computational markers (Figure 6 and Table 4).

**Figure 6.**
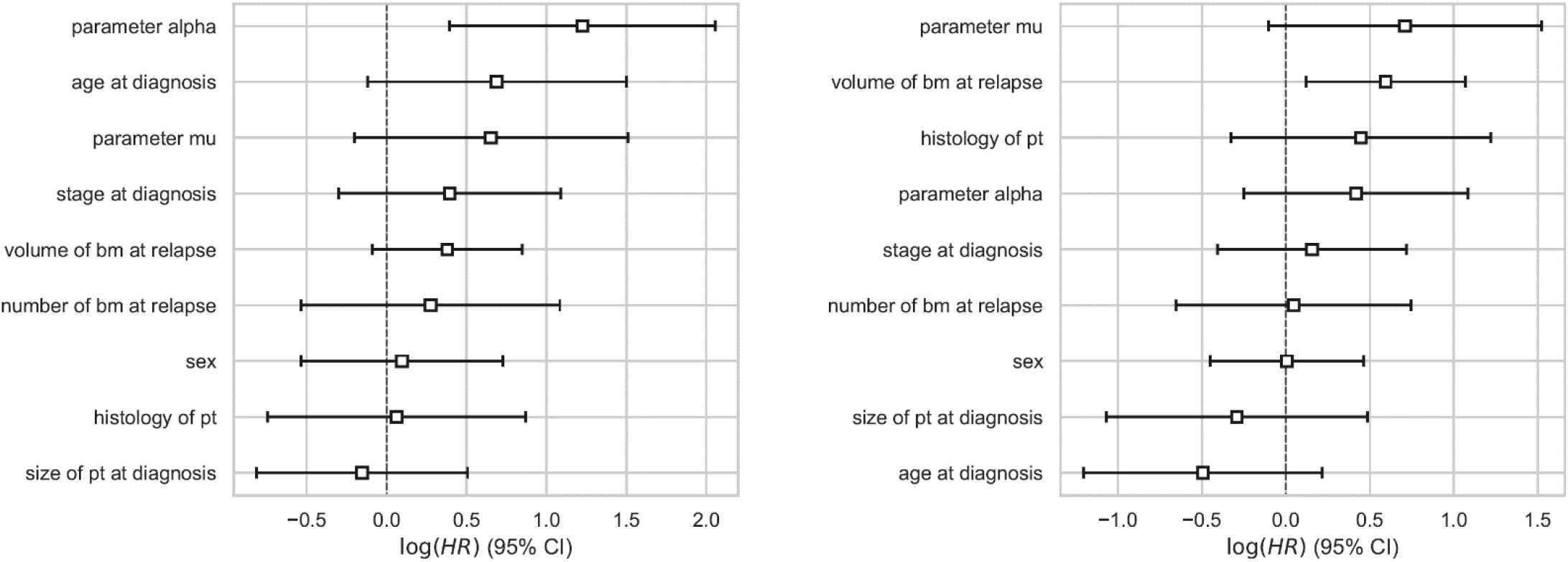
Hazard ratios for the predictive models. The hazard ratios were determined performing a multivariable Cox regression. The ratios are corresponding to the predictive models for OS (left) and PFS (right) with clinical and computational covariates, i.e., the settings where the determined parameters have been added to the respective patient data.

**Table 4.** Results of the multivariate Cox regression for OS and PFS. The table shows the characteristics of the results of the multivariate Cox regression for the predictive model including clinical and computational covariates for overall survival and progression-free survival. HR = hazard ratio, CI = confidence interval, BM = brain metastases, PT = primary tumor.

| Covariate | Hazard Ratio (OS) | p-value (OS) | 95% CI (OS) | Hazard Ratio (PFS) | p-value (PFS) | 95% CI (PFS) |
| --- | --- | --- | --- | --- | --- | --- |
| Sex | 1.1 | 0.769 | 0.584 - 2.07 | 1.01 | 0.979 | 0.637 - 1.59 |
| Age at diagnosis | 1.99 | 0.0951 | 0.887 - 4.47 | 0.61 | 0.174 | 0.3 - 1.24 |
| Number of bm at relapse | 1.32 | 0.506 | 0.586 - 2.96 | 1.05 | 0.894 | 0.52 - 2.11 |
| Stage at diagnosis | 1.48 | 0.266 | 0.74 - 2.97 | 1.17 | 0.585 | 0.667 - 2.05 |
| Histology of pt | 1.06 | 0.879 | 0.475 - 2.39 | 1.56 | 0.258 | 0.721 - 3.39 |
| Parameter $\alpha$ | 3.4 | 0.00387 | 1.48 - 7.81 | 1.52 | 0.219 | 0.78 - 2.96 |
| Parameter $\mu$ | 1.92 | 0.136 | 0.815 - 4.52 | 2.04 | 0.087 | 0.902 - 4.59 |
| Size of PT at diagnosis | 0.857 | 0.647 | 0.443 - 1.66 | 0.748 | 0.463 | 0.344 - 1.63 |
| Size of BM at relapse | 1.46 | 0.113 | 0.914 - 2.34 | 1.81 | 0.0143 | 1.13 - 2.91 |

For OS, the c-index for those two cases were 0.585 (95% CI 0.569 to 0.602) and 0.713 (95% CI 0.700 to 0.726), respectively (statistically significant increase, p<0.0001). This drastic increase of predictive value also held in reduced models containing only variables with p < 0.2 in the multivariate analysis (0.647 (95% CI 0.631 to 0.664) versus 0.789 (95% CI 0.779 to 0.800), statistically significant increase, p<0.0001). For PFS, the c-indices were 0.560 (95% CI 0.545 to 0.575) and 0.595 (95% CI 0.582 to 0.608), respectively (statistically significant increase, p<0.001). Considering again only variables with p < 0.2 in the multivariate regression yielded c-indices of 0.708 (95% CI 0.697 to 0.718) versus 0.703 (95% CI 0.693 to 0.713).

Interestingly in both cases for OS and PFS, the prediction model with clinical variables identifies the number of visible brain metastases as variable with large hazard ratio, but the prediction model with the computational markers selects *μ* as having better predictive power. That is, the number of brain metastases at first metastatic relapse loses significance when the prediction model is augmented with *μ*. For OS, the other selected covariates besides *μ* were *α*, the age at diagnosis and the summed volume of BMs at first metastatic relapse. For the PFS, these were the age at diagnosis and the summed volume of BMs at first metastatic relapse for PFS.

## DISCUSSION

Because of their prevalence and high morbidity, the management of BMs is a major issue in NSCLC. Given the heterogeneity of clinical cases and variability of treatments available, therapeutic decisions following intracranial oligoprogression are complex Predicting the onset and evolution of BMs in NSCLC patients using mathematical modeling would offer decision support with a personalized follow-up and treatment. The objective of this study was to test a mathematical model previously developed and to apply this model to a situation of intracranial oligoprogression in patients with early-stage NSCLC initially treated with a curative intent.

We first demonstrated that the mathematical model was suitable to describe clinical history of size and number of BMs in most patients. Simulations aligned with the data for 20 patients, while in eight cases the model underestimated the total number of BMs. However, among these eight cases the simulations correctly identified the presence of multiple BMs. Among the 8 patients, there are 3 squamous cell carcinomas and 5 adenocarcinomas, 3 of which had a mutation of interest: *KRAS G12C, STK11, BRAF*. These mutations may lead to a different growth of the primary tumor or a different metastatic dissemination. BRAF and STK11 have been described as genes involved in the metastatic process^69,70^. Moreover, one of the squamous cell carcinoma had a PTEN mutation, which has been shown to be associated with BM in breast cancer^71^. Epidermoid histology and molecular biology possibly misled the model.

In addition, predictive models of OS including the computational markers *α* and *μ* strongly outperformed models based on clinical variables only. For the metastatic growth rate *α*, significant differences were observed for OS and PFS. For the metastatic seeding *μ*, a significant difference was found at the 80% quantile for OS and PFS, thus identifying a fraction of high-risk patients.

Identification of a high-risk group for recurrence could assist the lung cancer physicians in therapeutic management.

In univariate analysis, WBRT at first relapse had a significant effect on PFS (HR 0.649 (95% CI 0.442 - 0.955), p=0.0282) and systemic treatment at first relapse had a minor non-significant effect (HR 0.824 (95% CI 0.546 – 1.25), p=0.359). This could be due to undetected brain metastases at *T*_*BM*_, or dormant metastatic cells. Systemic treatment or WBRT may therefore be of value for patients at risk. Indeed, WBRT has the advantage of covering all macroscopic and microscopic BM to control cerebral progression. However, WBRT does not increase survival^21,22^ and leads to neurocognitive decline^23,24^. The effect of systemic treatments seems less obvious given their variable intracranial efficacy. Our model could lead to a computational tool that could assist lung cancer clinical oncologists in identifying patients with high-risk of multiple recurrences that could benefit from systemic treatments.

Other tools exist to help the management of NSCLC patients. Some are based on molecular biology prognostic scores such as the Lung-Mol Score^40^, others rely on radiomics features^72–75^. For example, Huang et al. demonstrated that radiomics features extracted from pre-treatment T1 MRI was an independent prognostic factor of local control for BM patients who underwent gamma knife radiosurgery^74^. The advantage of our mathematical model compared to these tools is, that it is of biologically grounded and mechanistic nature. In turn, this allows to perform simulations of the disease and clinical course combined with predictive power, unlike the prognostic tools mentioned above.

The main limitation of our study is the number of patients included. It will be necessary to confirm our findings on a larger number of patients and to include molecular profiling data. In addition, we could conduct a prospective study to keep double-blinded design for predictions.

On the other hand, the population chosen in this study (patients with early-stage NSCLC initially treated with a curative intent and presenting with intracranial oligoprogression) limits the application of our results. However, this made it possible to limit bias of systemic treatments and in this situation, lung cancer physicians are facing an unmet need with no existing guidelines.

The modeling approach remains a simplified description of the complexity of the metastatic cascade that is yet incompletely understood^76^. However, considering the limited data available, the model complexity had to be reduced to ensure practical identifiability of the parameters. This resulted in only two critical free parameters controlling growth (*α*) and dissemination (*μ*). An assumption that leaves for improvement is that primary tumor growth parameters were determined from meta-analyses values using only the tumor pathology^60^. We need further investigation to understand the influence of molecular biology, especially EGFR mutations^77,78^ or ALK translocation because these patients have more multiple brain relapses but have better prognosis^79^. Moreover we need to understand the impact of treatments on the primary tumor, particularly with the advent of neoadjuvant and adjuvant treatments, which could reduce micrometastatic disease and thus risk of relapse^80–82^.Moreover, our model could be extended to account for more complex processes of the metastatic cascade, such as dormancy^83^ and the possibility that the primary tumor is not the only one that can spread new lesions^84^. Regarding the relapse site, we must understand the influence of the blood-brain barrier and the unique brain microenvironment^85^.

However, simplifying the metastatic cascade increases the feasibility of the use of this model in clinical practice. Once the model is validated on a larger scale, applications will be developed for the clinicians to determine the risk of cerebral recurrence for each patient by selecting two or three clinical parameters available in routine.

To our knowledge, this is the first mechanistic modeling approach to predict the occurrence and growth of BMs in NSCLC with data routinely acquired in the clinics. This work could allow to identify patients at high risk of recurrence and to propose a personalized management such as follow-up imaging schedule, addition of systemic or non-systemic therapy, and use of WBRT depending on the patient’s risk of multiple recurrences.

## Data Availability

The datasets used and analyzed during this study are available from the corresponding author on reasonable request.

## ACKNOWLEDGEMENTS

PS wants to thank Christina Kuttler for valuable discussions on the mathematical modelling method. Supported by Deutsche Forschungsgemeinschaft (DFG) through TUM International Graduate School of Science and Engineering (IGSSE), GSC 81.

## CONTRIBUTIONS

Conceived the research idea: SB, PT. Model setup: PS, SB. Collected the data: ES, PT. Performed data analysis and presentation: PS. Wrote software to estimate parameters and simulate: PS, SB. The paper was written by ES and PS with editorial input from all authors. All authors read and approved the final manuscript.

## SUPPLEMENTARY MATERIAL

### S1: Analytical solution of the primary tumor and metastases’ growth dynamics

Variables and parameters are explained in the text. The analytical solution to the primary tumor dynamics can be determined to be

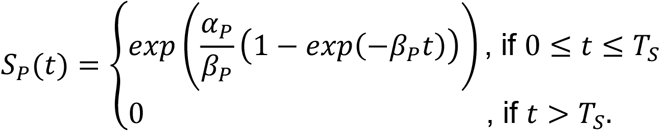

and the metastases’ Gompertz growth function is given analogously, without the possible surgery modelled, as

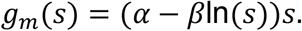

The metastatic Gompertzian growth equation as analytical solution of the equation 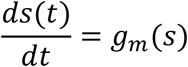 with initial condition *s*(0) = 1 that describes the metastatic size *s* at time *t* ≥ 0 reads

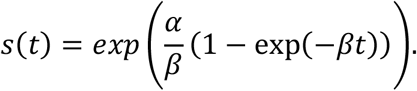

### S2: Model equation for the metastatic density ρ(s, t) of size s and time t

The full equation corresponding to the approach of ^60^ modelling the metastatic density *ρ*(*s, t*) as function of size *s* and time *t* with boundary and initial conditions neglecting secondary metastasation – not considering the integral in the boundary condition – reads

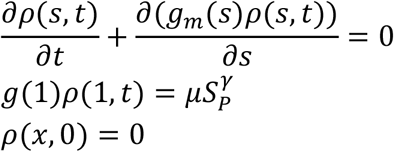

The used parameters are explained in the text.

### S3: Parameter estimation: growth parameters

The doubling time *τ*_*P*_(*s*) of a primary tumor of size *S*_*P*_(*T*_*d*_) at time of diagnosis 0 ≤ *T*_*d*_ ≤

*T*_*S*_, i.e. the time a tumor of size *s* needs to grow to a size of 2*s*, is implicitly given by^51^

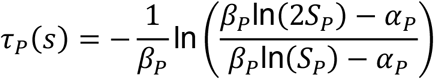

and from *exp*(*α*_*P*_/*β*_*P*_) = 10^12^ we determined the parameters *α*_*P*_ and *β*_*P*_ from the primary tumor histology and the primary tumor size at diagnosis. Since on the other hand *exp*(*α*/*β*) = 10^12^ for the metastases’ dynamics, the parameter *β* was determined from the value of parameter *α*, which in turn was a fitted parameter (cf. Fitting procedure).

### S4: Numerical implementation: calculate metastases

Since we assumed that the primary tumor was the only source for metastases, we applied the procedure from ^60^ to calculate the absolute number of metastases *N*(*t*) at a certain time *t* from

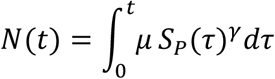

and to introduce the cumulative size distribution *f*(*t, s*) as the number of metastases larger than a fixed size *s* at a certain time *t* as

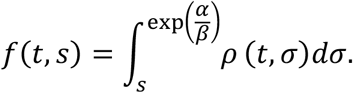

With *t*(*s*) as the time for a tumor to reach size *s* under Gompertz growth^60^ we receive

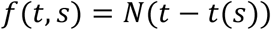

With

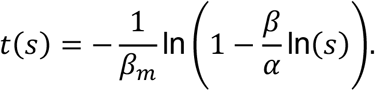

These numbers are the values that we observed clinically - the visible BMs in the data are assumed to correspond to the sizes of metastases larger than the volume corresponding to a diameter measurement of three mm. Thus, the number of metastases larger than this threshold size *s*_*v*_ at time *t* can be calculated as *f*(*t, s*_*v*_).

### S5: Model simulations for every individual patient of the data set

The explanation of every subpanel is given in the description of figure 3 of the main text.

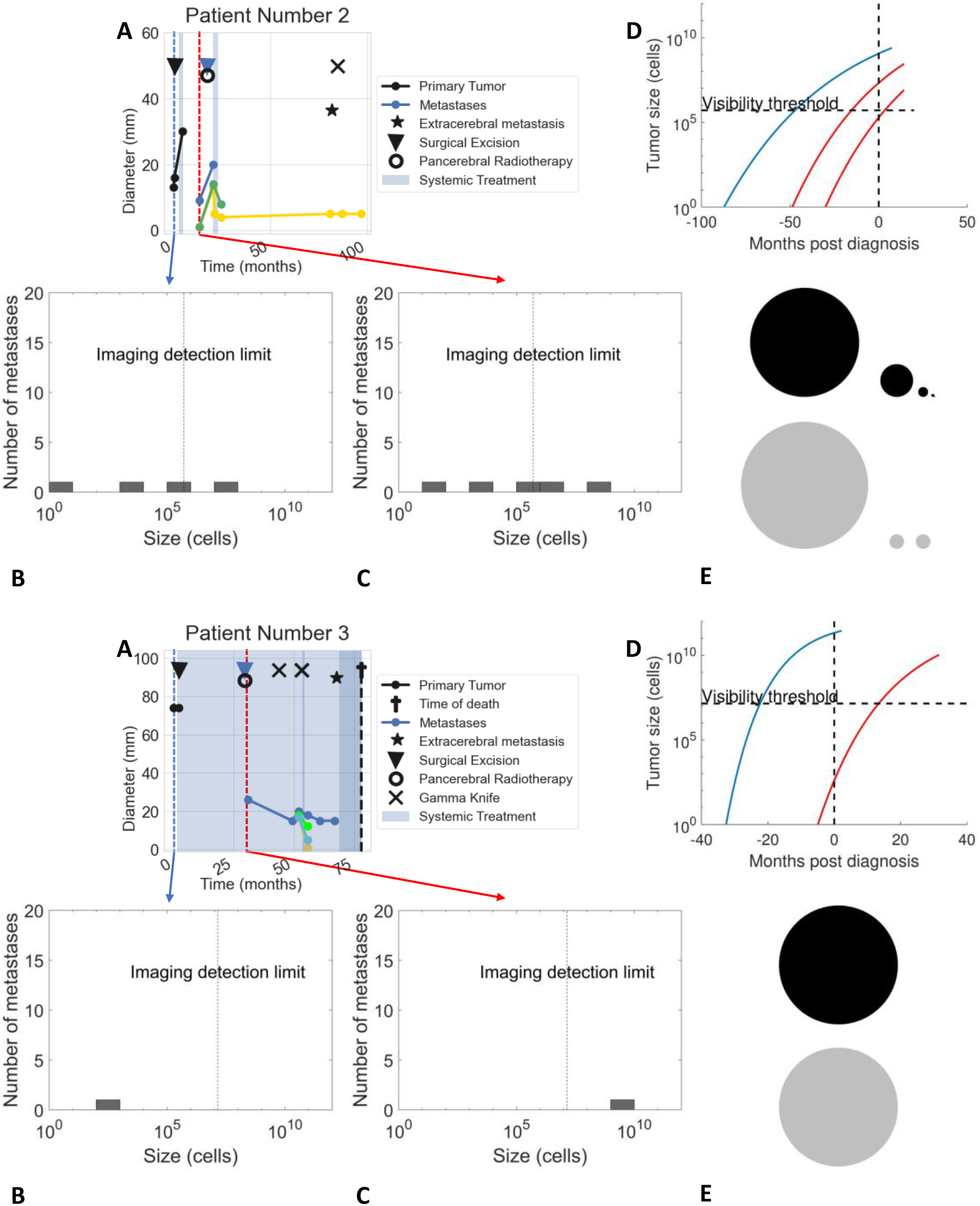

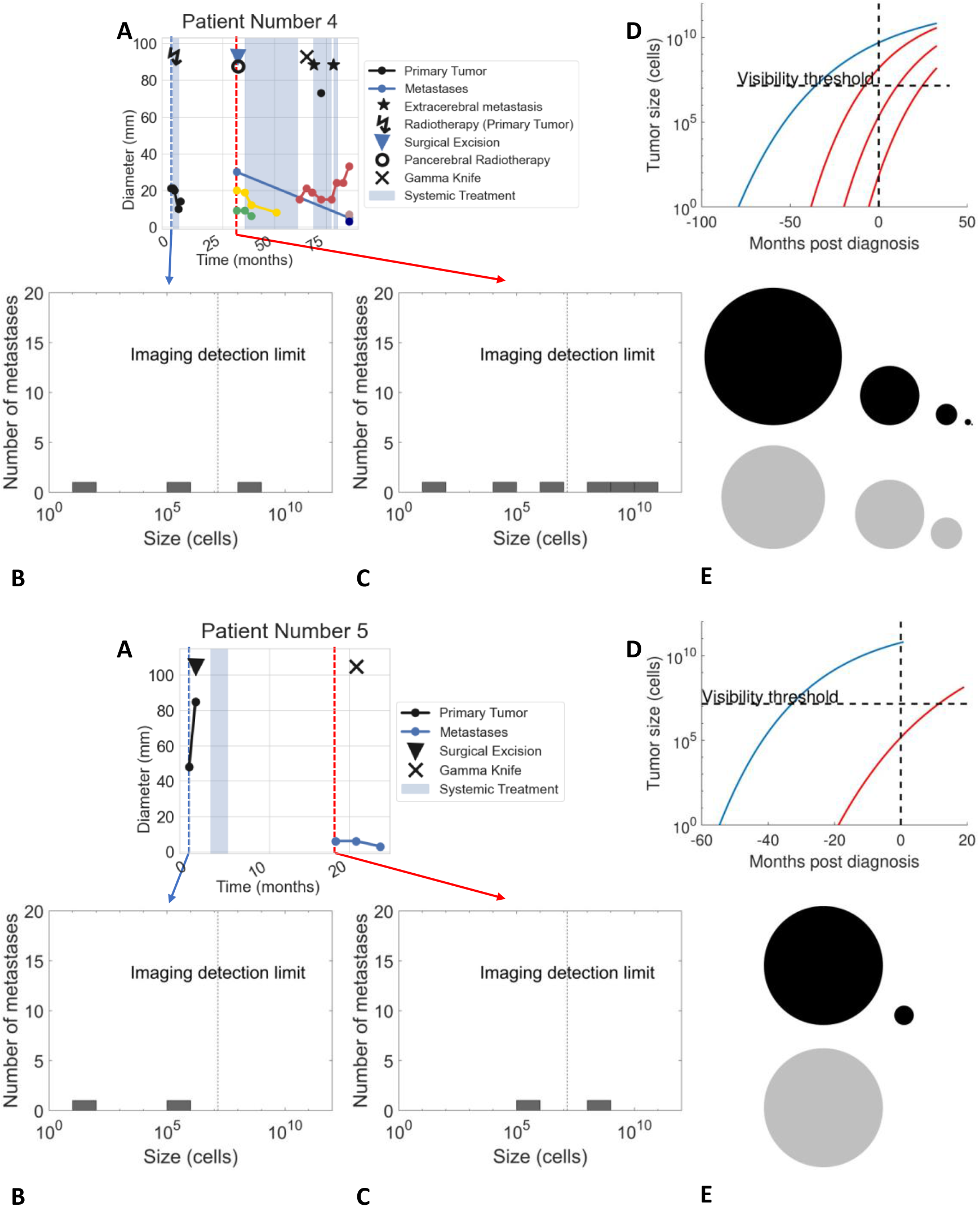

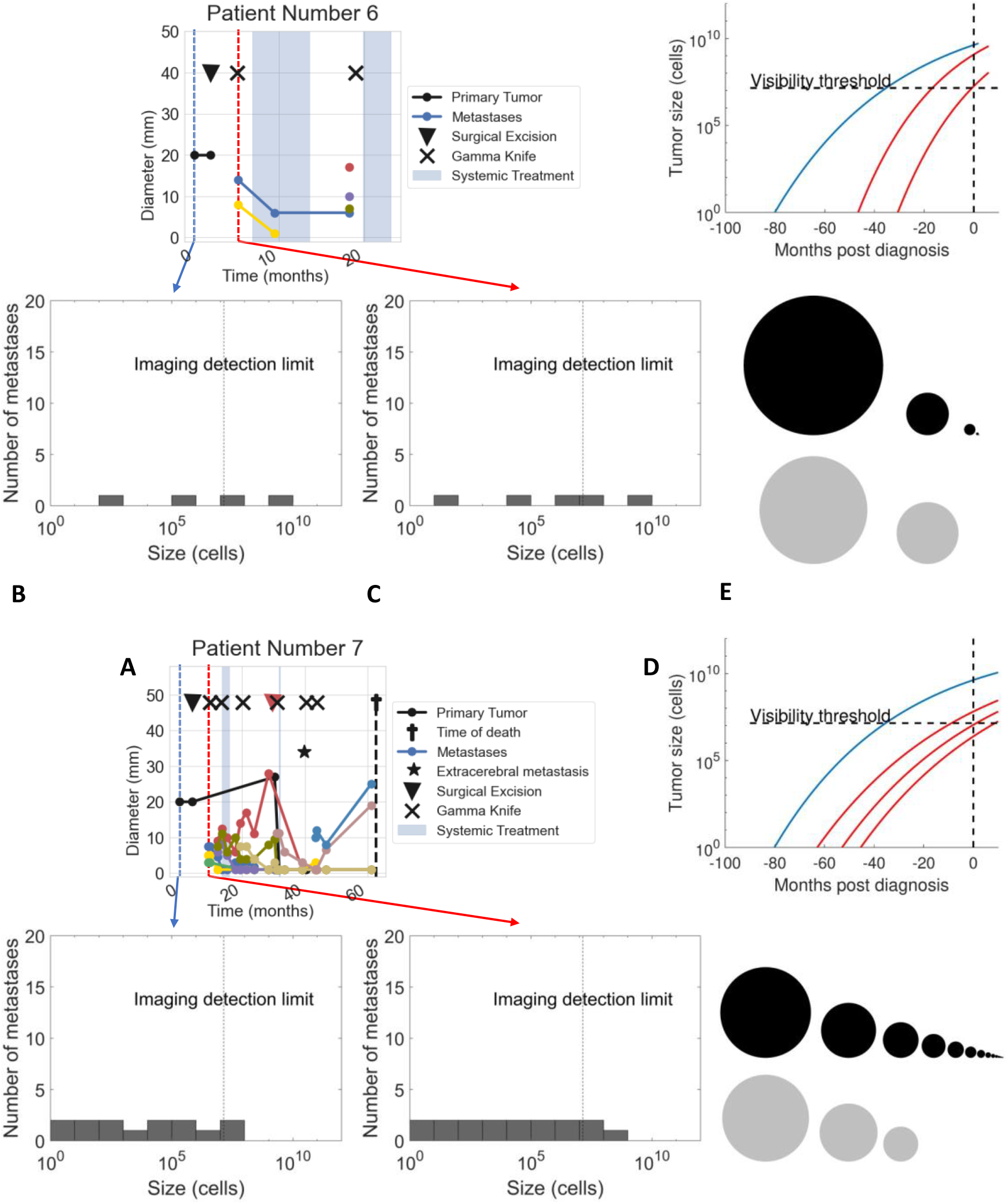

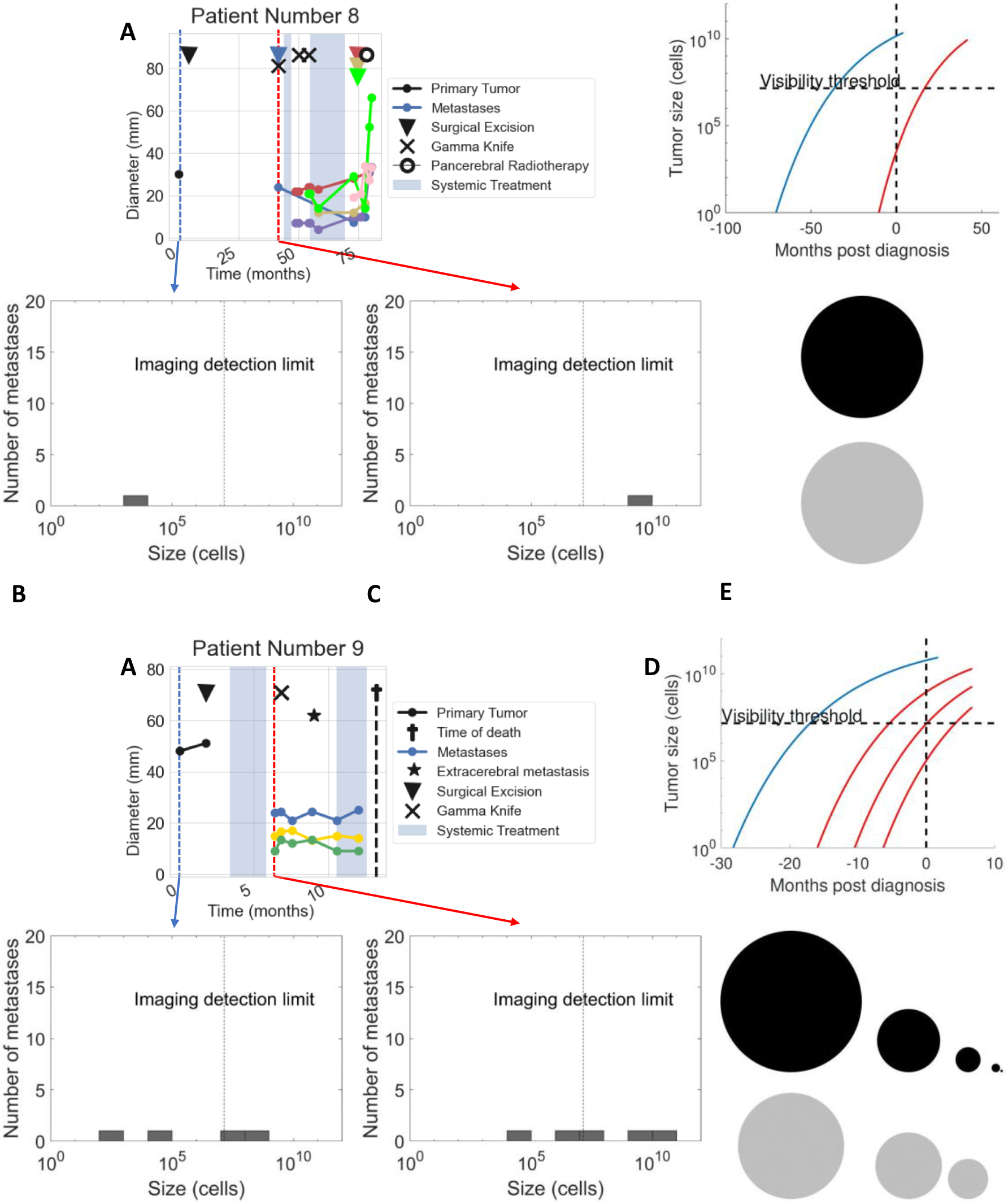

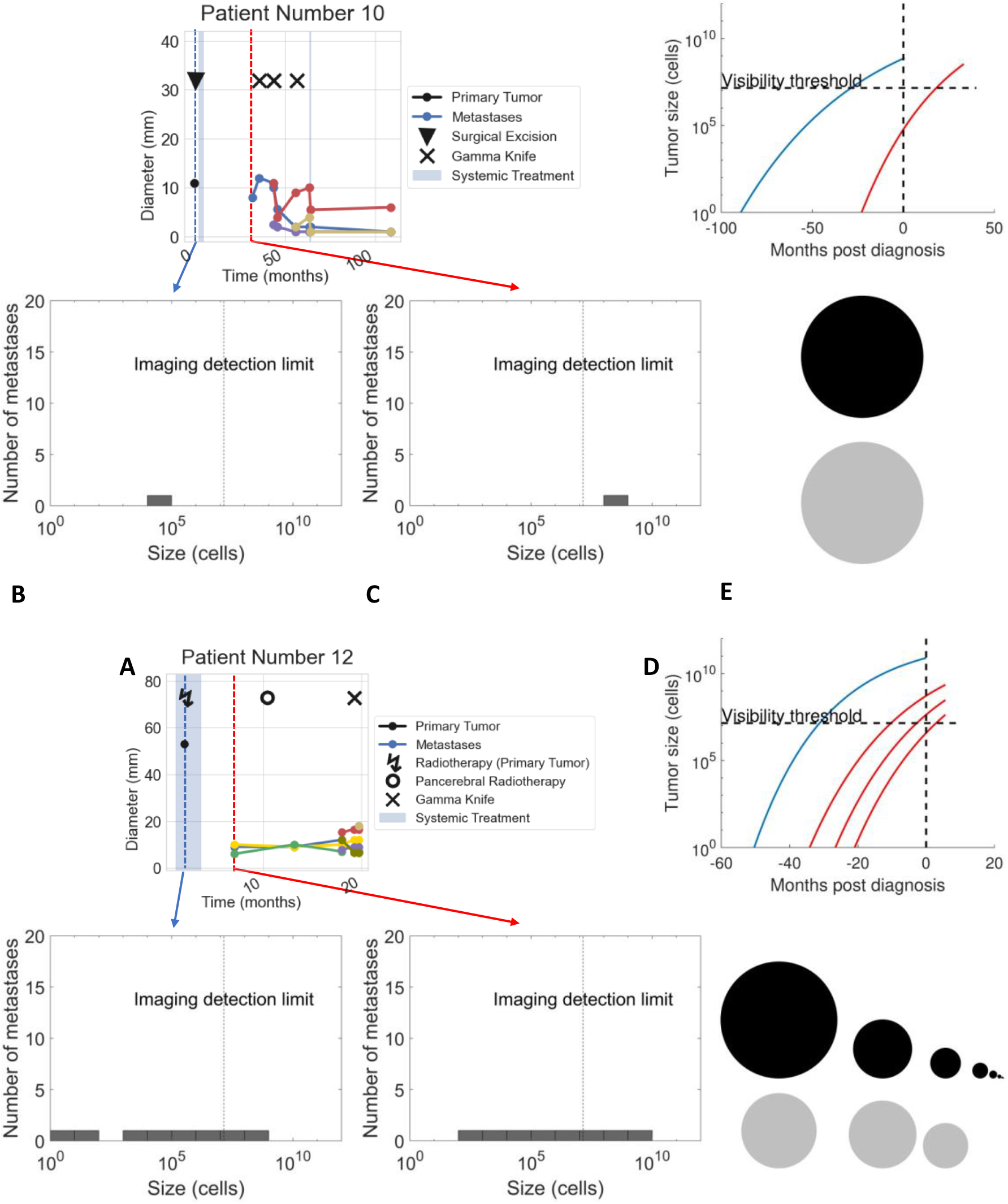

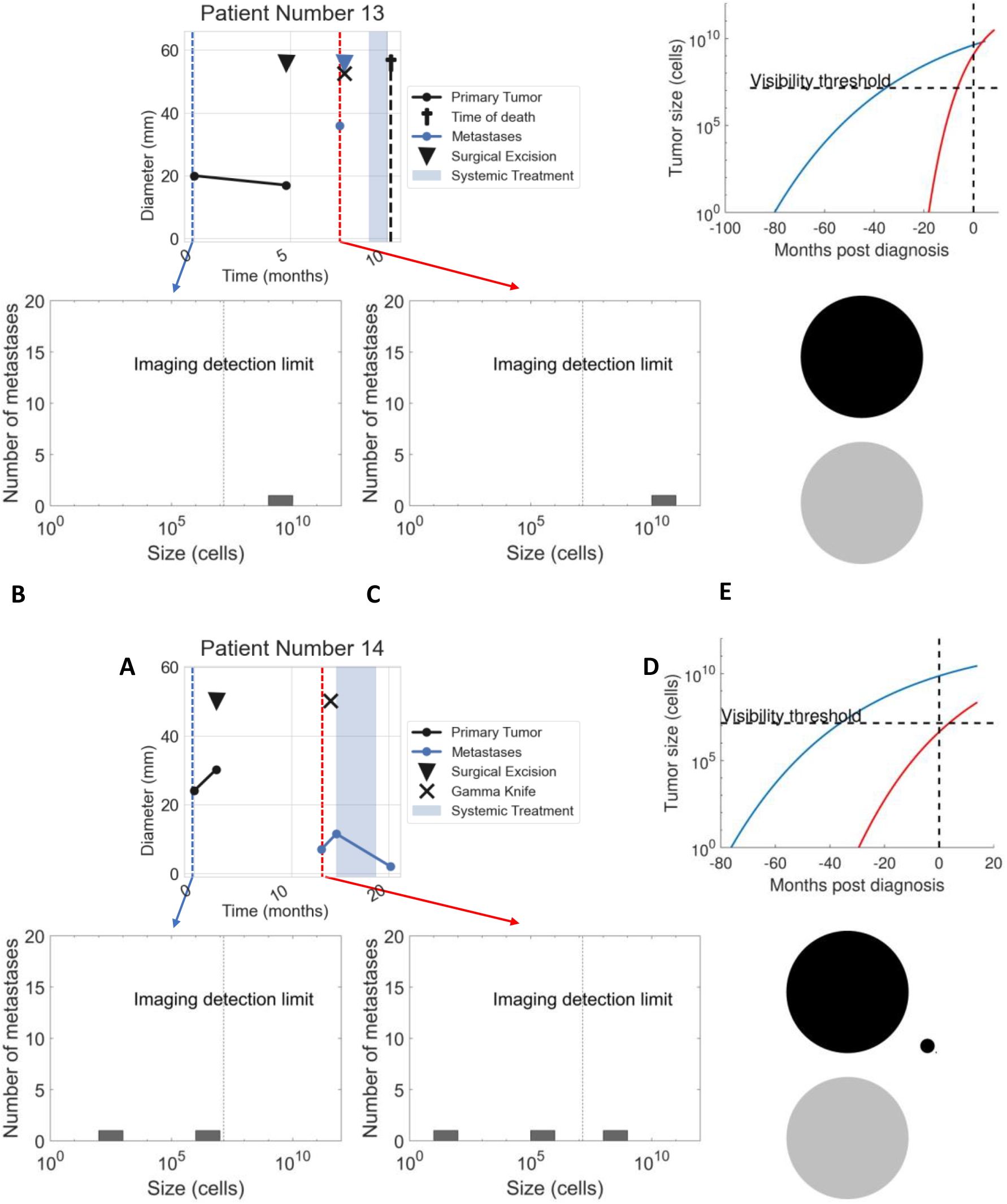

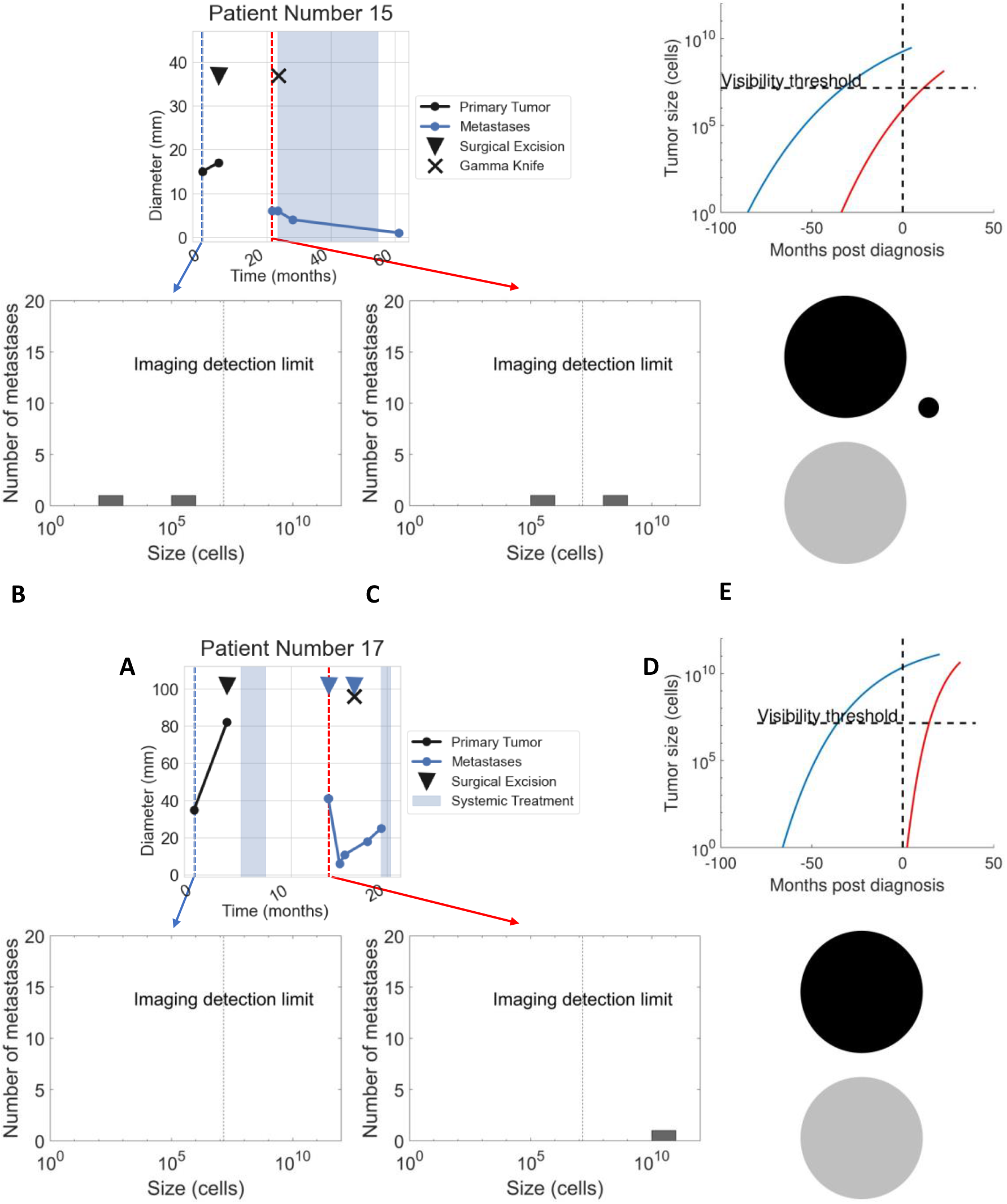

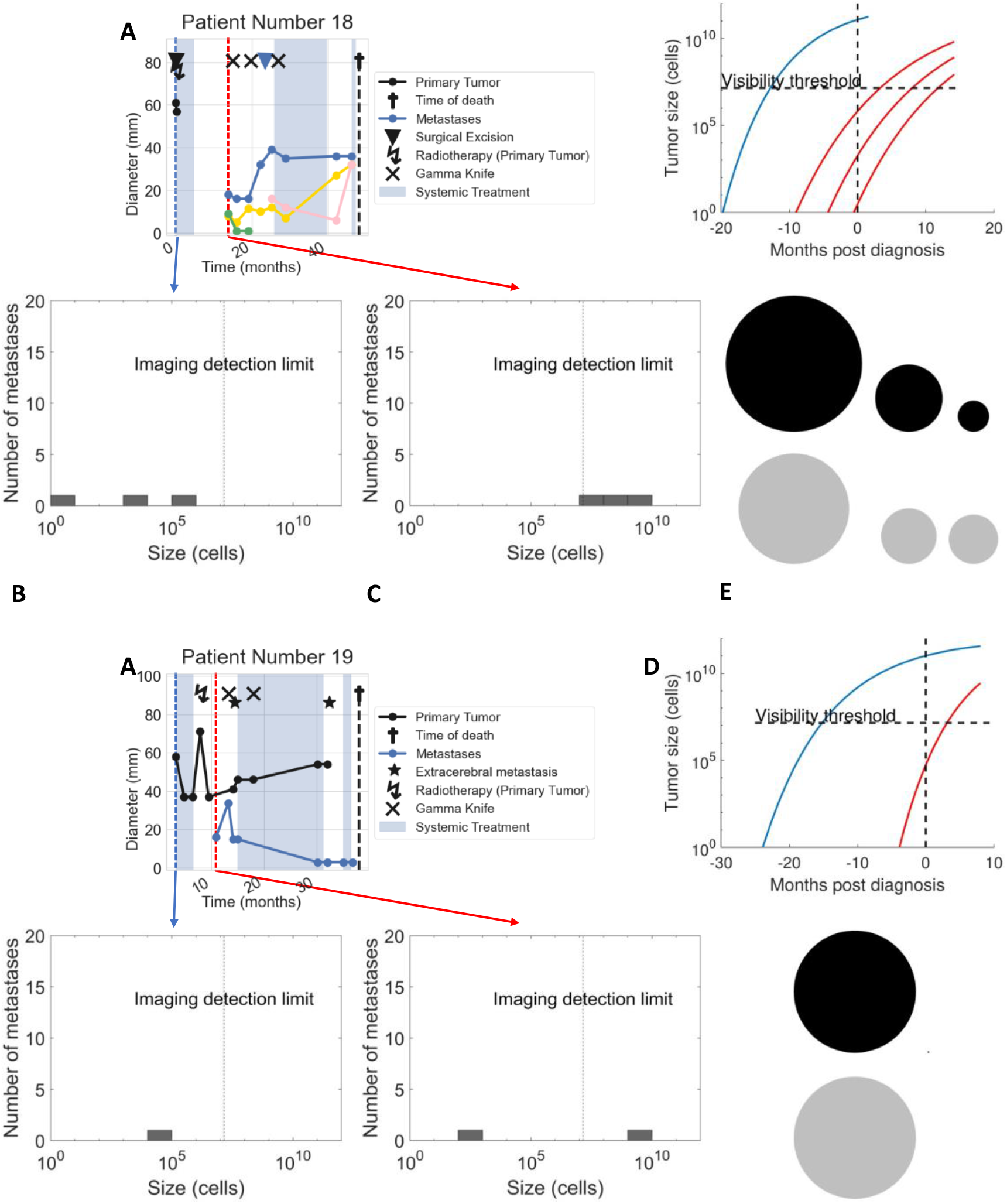

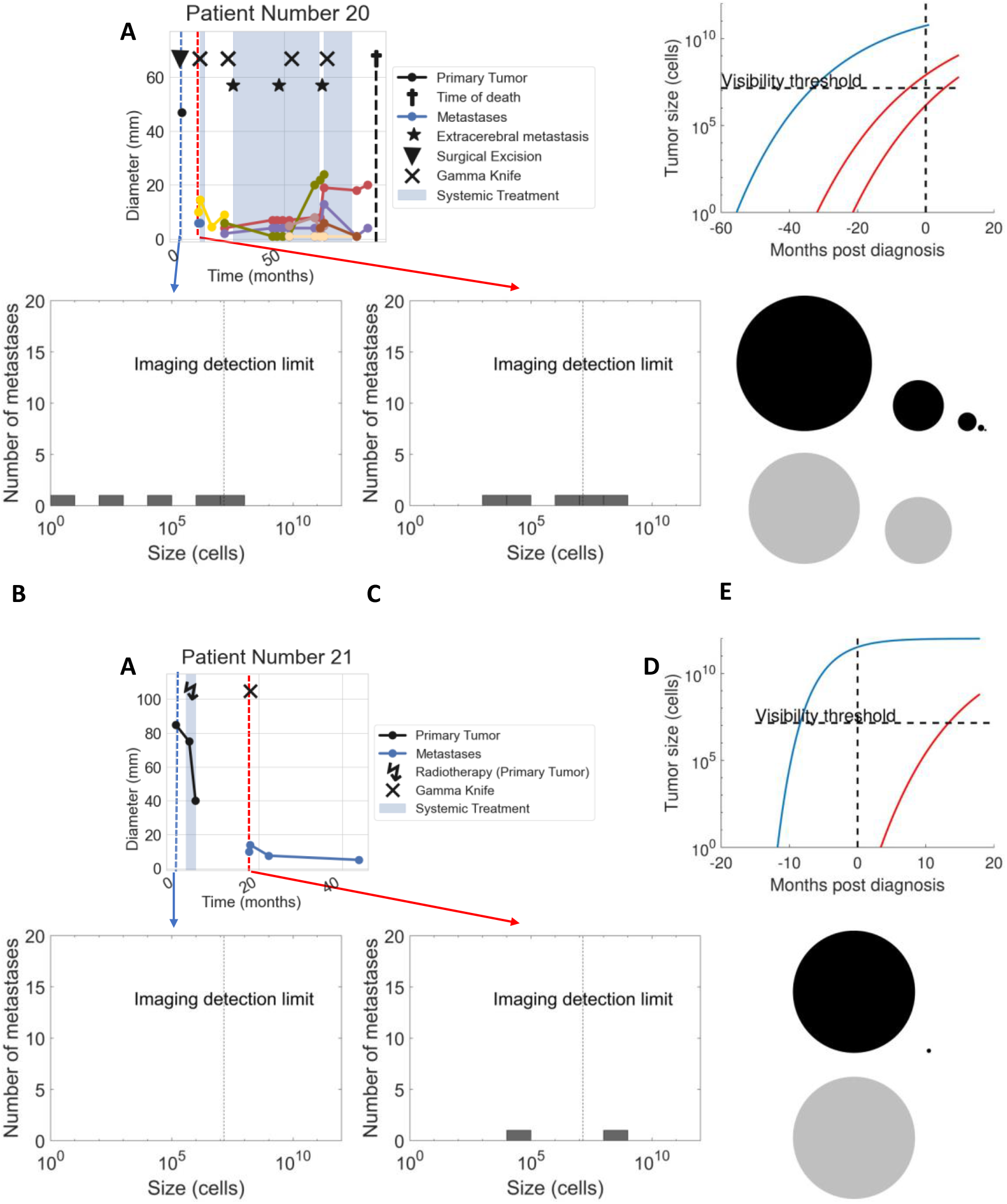

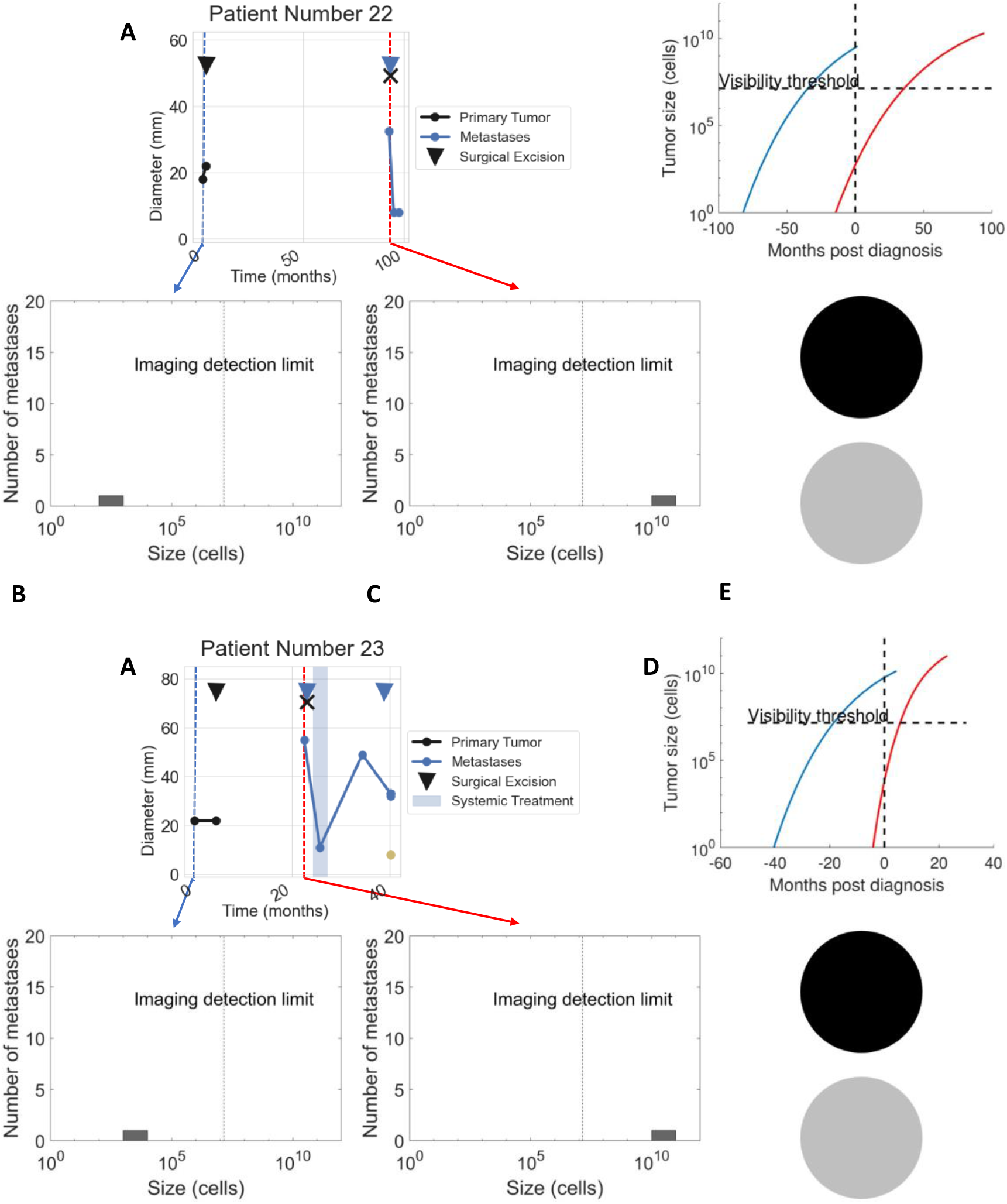

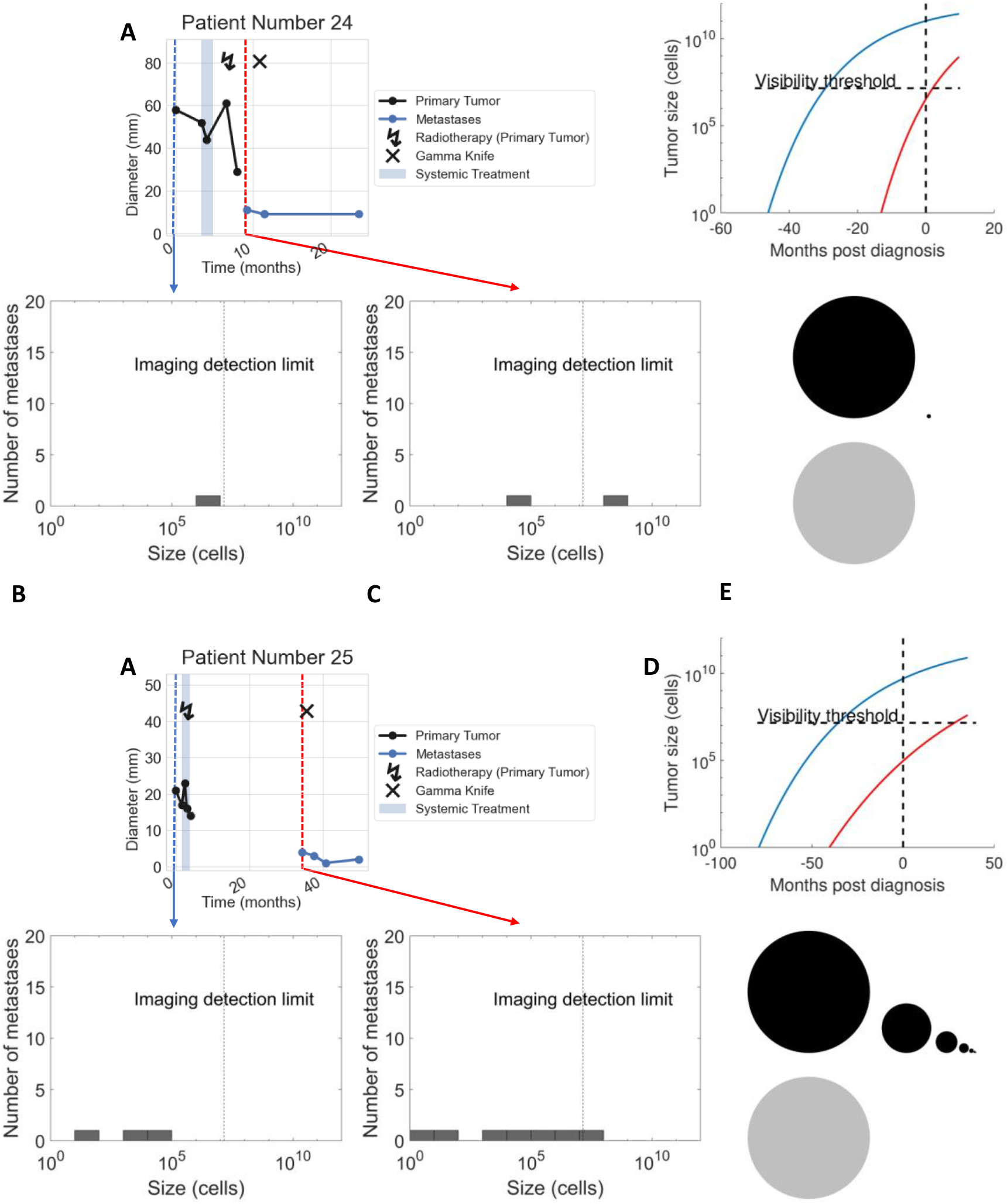

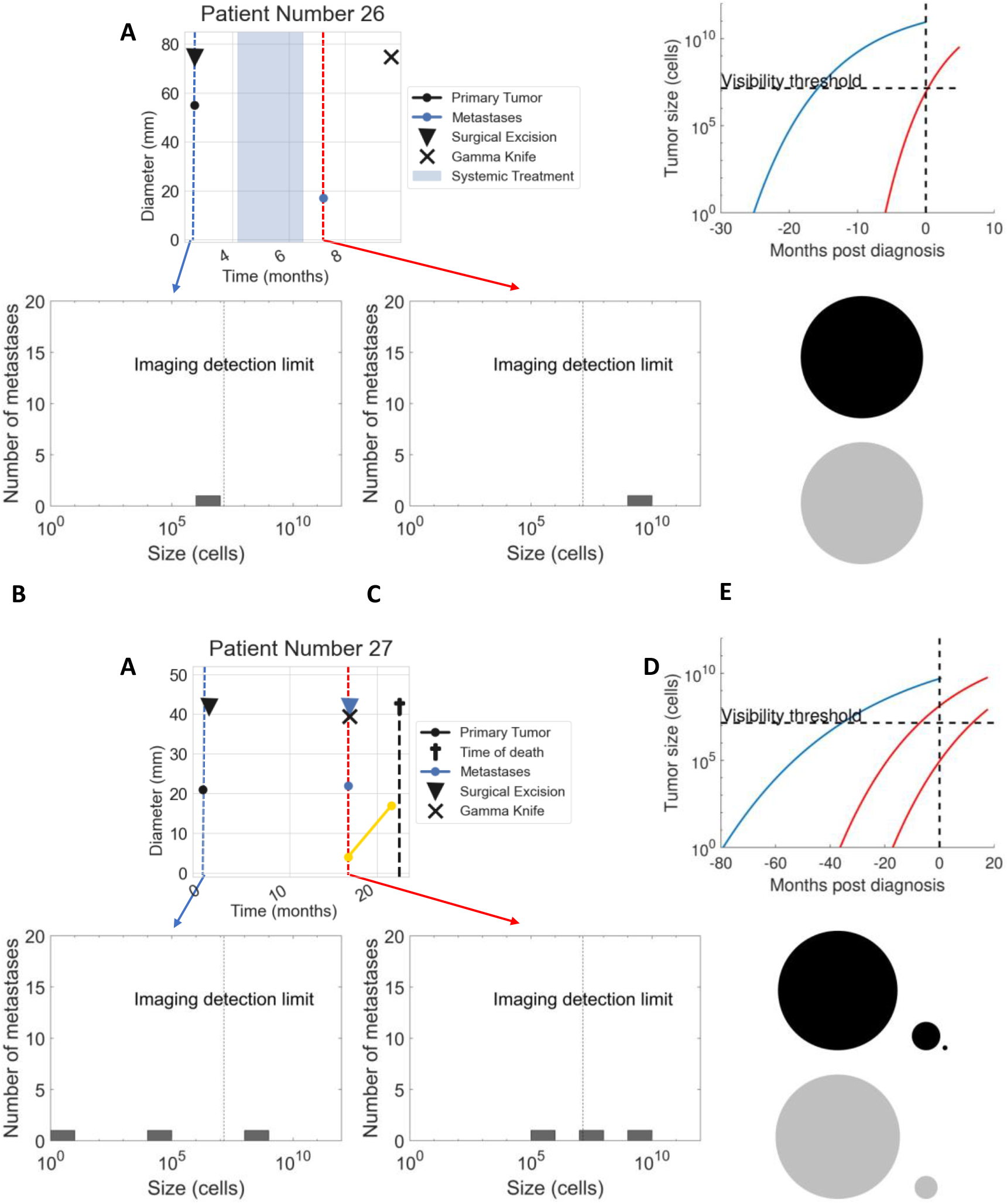

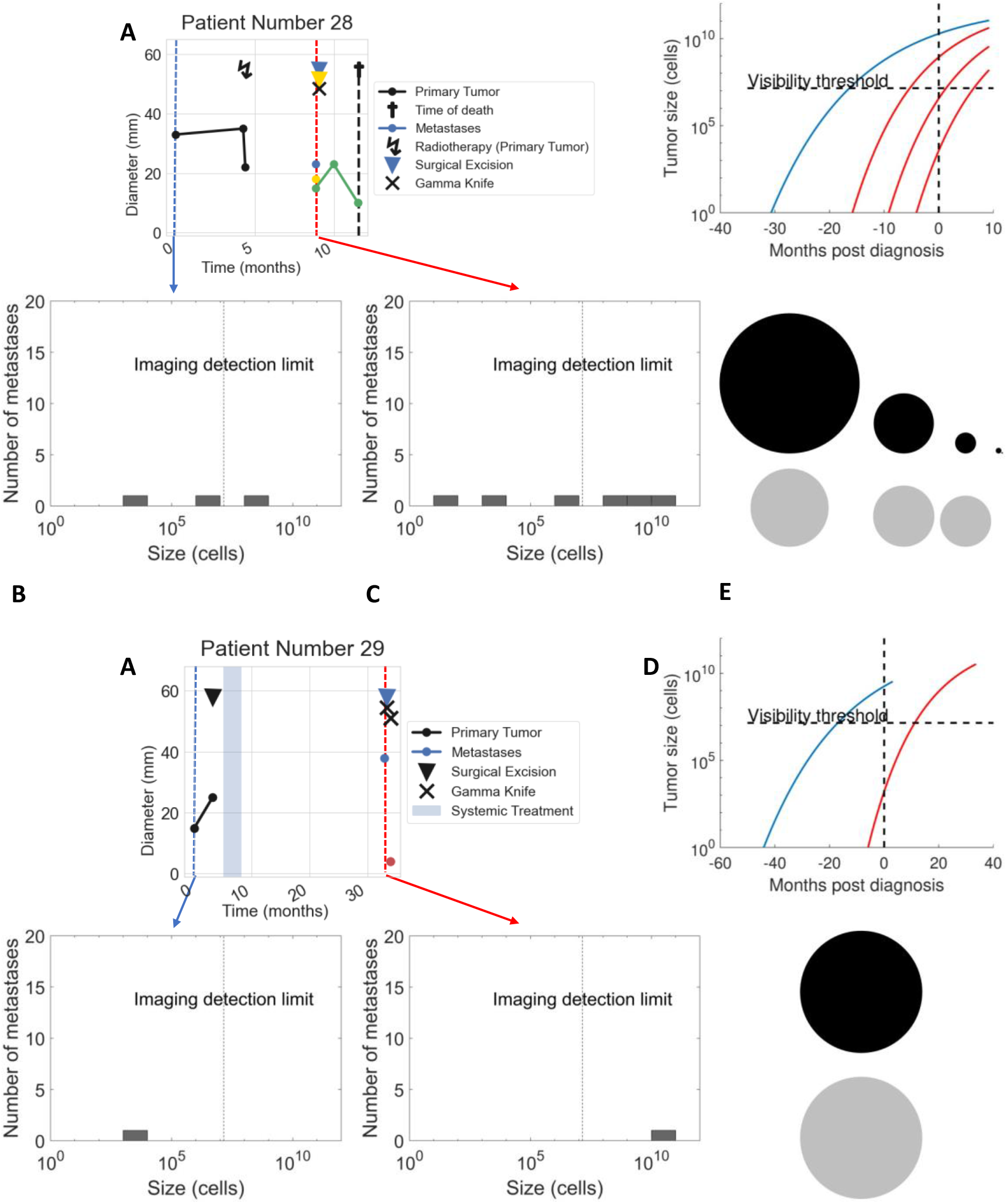

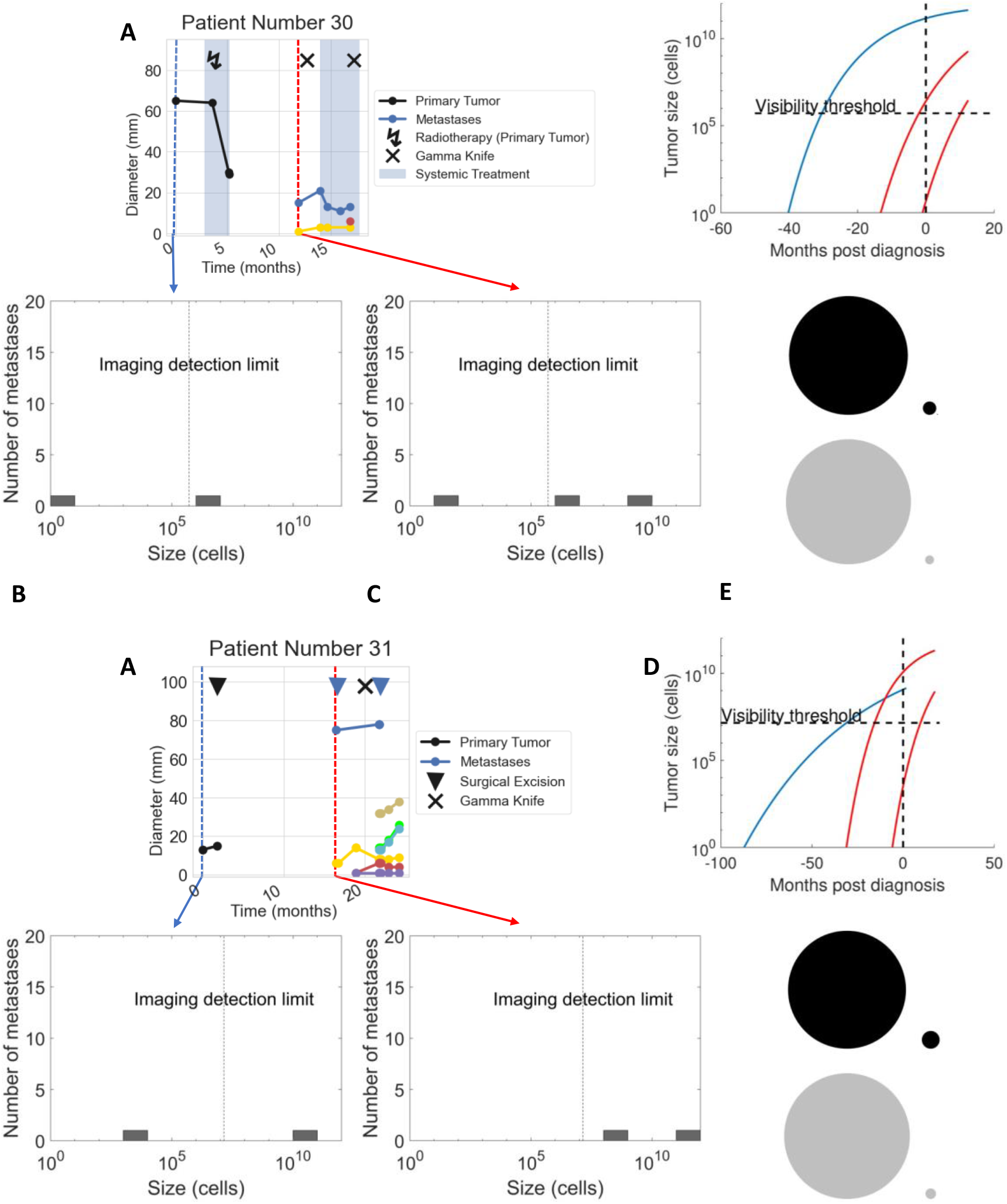

